# Performance of VIDAS® Diagnostic Tests for the Automated Detection of Dengue Virus NS1 Antigen and of Anti-Dengue Virus IgM and IgG Antibodies: a Multicentre, International Study

**DOI:** 10.1101/2022.03.16.22272491

**Authors:** Alice F. Versiani, Antoinette Kaboré, Ludovic Brossault, Loïc Dromenq, Thayza M.I.L. dos Santos, Bruno H.G.A. Milhim, Cássia F. Estofolete, Assana Cissé, Abel C. Sorgho, Florence Senot, Marie Tessonneau, Serge Diagbouga, Mauricio L. Nogueira

## Abstract

Dengue is a serious mosquito-transmitted disease caused by the dengue virus (DENV). Rapid and reliable diagnosis of DENV infection is urgently needed in dengue-endemic regions. We describe here the performance evaluation of the CE-marked VIDAS^®^ dengue immunoassays developed for the automated detection of DENV NS1 antigen, and anti-DENV IgM and IgG antibodies. A multicenter concordance study was conducted in 1296 patients from dengue-endemic regions in Asia, Latin America, and Africa. VIDAS^®^ dengue results were compared to those of competitor enzyme-linked immunosorbent assays (ELISA). The VIDAS^®^ dengue assays showed high precision (CV ≤ 10.7%) and limited cross-reactivity (≤ 15.4%) with other infections. VIDAS^®^ DENGUE NS1 Ag showed high positive and negative percent agreement (92.8% PPA, 91.7% NPA) in acute patients within 0–5 days of symptom onset. VIDAS^®^ Anti-DENGUE IgM and IgG showed a moderate to high concordance with ELISA (74.8% to 90.6%) in post-acute and recovery patients. PPA was further improved in combined VIDAS^®^ NS1/IgM (96.4% in 0–5 days acute patients) and IgM/IgG (91.9% in post-acute patients) tests. Altogether, the VIDAS^®^ dengue NS1, IgM and IgG assays performed well, either alone or in combination, and should be suitable for the accurate diagnosis of DENV infection in dengue-endemic regions.

## 1. Introduction

Dengue is a mosquito-transmitted disease caused by one of the four dengue virus (DENV) serotype (DENV-1 to DENV-4) [1,2]. With an estimated 96 million annual symptomatic infections, spreading mainly in Asia (70%), Africa (16%) and the Americas (14%), dengue disease is a major public health concern [2–5]. The course of the disease ranges from mild to severe, with an early febrile (acute) phase lasting 2–7 days followed by a critical phase that may either worsen, evolving to a life-threatening course (severe dengue), or improve to full recovery usually within 1–2 weeks [1,2,5]. In the absence of specific antiviral therapy, supporting care can effectively control severe dengue progression [1,2,5]. Administration of supportive care and other management protocols highly depend on early and reliable dengue diagnosis. Early dengue diagnosis based on clinical manifestations is challenging, as early symptoms do not differentiate between dengue and other febrile diseases [1,2,5,6]. Beside individual patient management, reliable and rapid DENV infection diagnosis is crucial to monitor and control dengue outbreaks in dengue-endemic regions [1,2,5,6]. Therefore, there is an urgent need for easy-to-use, rapid and accurate DENV-specific diagnostic assays.

Current guidelines recommend detecting circulating DENV RNA by reverse-transcription PCR (RT-PCR) and/or the secreted viral non-structural protein 1 (NS1) antigen by immunoassay within the first 5–7 days of illness (acute phase) to confirm DENV infection [5,7,8]. Following the acute phase, the detection of anti-DENV immunoglobulin M (IgM) and/or G (IgG) using an immunoassay, such as enzyme-linked immunosorbent assay (ELISA) or immuno-chromatographic rapid diagnostic test (RDT), is highly suggestive of a prior (primary or secondary) DENV infection [5,7,8]. Because of the kinetics of response of these different markers (DENV RNA, DENV NS1 antigen, anti-DENV IgM and IgG), their combined testing can increase the sensitivity of diagnosis, by extending the DENV diagnostic window [1,2,5,7–16].

RDT have been widely implemented in field settings. They are easy to use and rapid (about 20 minutes to interpretation). Their visual interpretation is however operator-dependent [17] and they lack in sensitivity [9,12,16,18–23]. ELISA are more sensitive than RDT [9,12,16,18–23] but their manual execution limits their routine implementation in field settings, notably at time of dengue outbreaks. We recently described the performance of three VIDAS^®^ dengue prototype assays developed for the detection of DENV NS1 antigen and anti-DENV IgM and IgG antibodies [24]. VIDAS^®^ dengue are fully automated immunoassays intended as an aid in the diagnosis of DENV infection in symptomatic patients. The three VIDAS^®^ dengue assays can be tested in parallel or applied independently. They are rapid (40– 60 minutes to result), easy to use, and easy to interpret (positive or negative) with no equivocal zone. Prototype performance was evaluated in 91 Lao patients with acute DENV infection. We showed that VIDAS^®^ dengue prototypes performed well, in comparison to manual competitor ELISA, both in adults and children, and that they outperformed RDT in the diagnosis of acute DENV infection [24].

The aim of this international, multicenter study was to evaluate the clinical performance of the three CE-marked VIDAS^®^ dengue immunoassays VIDAS^®^ DENGUE NS1 Ag (DEAG), VIDAS^®^ Anti-DENGUE IgM (DENM) and VIDAS^®^ Anti-DENGUE IgG (DENG) in the main dengue-endemic regions of the world (Asia, Latin America and Africa). A total of 1296 samples of patients with suspected DENV infection were tested with the VIDAS^®^ dengue assays and compared to manual competitor ELISA. Results were interpreted using a stage-based algorithm considering the outcome of the three dengue NS1, IgM and IgG assays, and defining different stages of DENV infection (acute, post-acute and recovery) as well as a naïve status (i.e., patients with other febrile illnesses). Furthermore, we evaluated the analytical performance (precision, cross-reactivity) of the three VIDAS^®^ dengue immunoassays. Altogether, this multicenter study demonstrated that the VIDAS^®^ dengue NS1, IgM and IgG assays performed well, either alone or in combination, and therefore that they should be suitable for the accurate diagnosis of DENV infection in febrile patients of dengue-endemic regions.

## 2. Materials and Methods

### 2.1. Patients and Samples

A total of 1636 sera were collected between September 2015 and October 2020 in patients with a suspected DENV infection, presenting at the hospital with one or more of the following symptoms: fever, headache, diarrhea, myalgia, arthralgia, retro-orbital pain, thrombocytopenia, edema, rash, and nausea (Figure 1). Samples were collected prospectively or retrospectively in several dengue-endemic regions, including Asia (India, Vietnam, The Philippines), Latin America (Brazil, Peru, Honduras, Dominican Republic) and Africa (Burkina Faso) and tested in central laboratories at three distinct sites, as described in Table 1.

**Figure 1.**
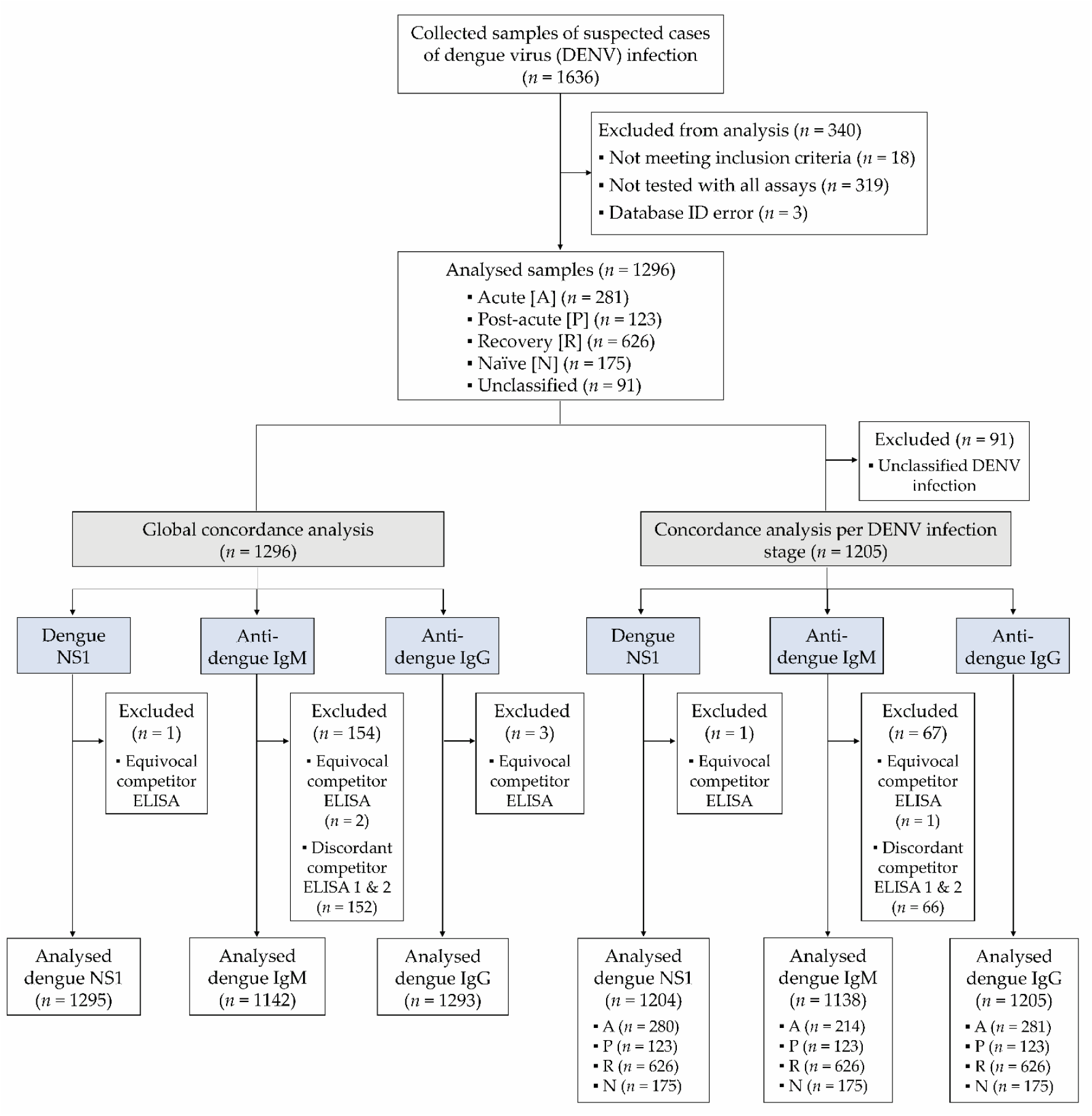
Study Flow diagram. A total of 1636 samples of suspected DENV infection were collected, of which 1296 eligible samples tested with all assays (competitor ELISA, VIDAS^®^ and PCR) were included in the analysis. Concordance analyses were conducted in the total population (*n* = 1296) and according to the DENV infection stages defined in Table 3 (*n* = 1205). An additional analysis on samples common to all immunoassays (*n* = 1138) is shown in Figure S2 and Table S5.

**Table 1.** Study samples

| Site | Collection site | Samples | Collection time | Testing site |
| --- | --- | --- | --- | --- |
| 1 | Hôpital Sainte Camille & Centre de Recherche Biomoléculaire Pietro Annigoni (CERBA), Ouagadougou, Burkina Faso | Prospective cohort Adults and children | June 2020 – October 2020 | Institut de Recherche en Sciences de la Santé (IRSS), Ouagadougou, Burkina Faso |
| 2 | Centro Integrado de Pesquisa, Hospital de Base, São José do Rio Preto, Brazil | Retrospective & prospective cohorts <sup>1</sup><br>Adults | February 2019 – August 2020 | Laboratório de Pesquisas em Virologia, FAMERP, São José do Rio Preto, Brazil |
| 3 | India, Vietnam, The Philippines, Peru, Honduras, and Dominican Republic | Retrospective cohort <sup>2</sup><br>Adults and children | September 2015 – September 2020 | Clinical Affairs Laboratory, bioMérieux, Marcy l'Etoile, France |

<sup>1</sup> Due to the COVID-19 pandemic, only two samples were collected prospectively; most samples were from a retrospective cohort collected during the 2019 dengue epidemic outbreak in Brazil. <sup>2</sup> Samples purchased from Boca Biolistics (Pompano Beach, Florida, USA), Discovery Life Sciences (Huntsville, Alabama, USA) and Atreide Biosamples (Hannover, Germany).

All collected sera (≥ 1.5 ml) were aliquoted to allow testing with the different assays on the same freeze/thaw cycle. Aliquots were frozen at the collection site, transported frozen under controlled conditions, and stored frozen at -20°C until testing. This study was conducted in adherence to the guidelines of the Declaration of Helsinki, and approved by the respective institutional review board at each institution (Comité d’Éthique pour la Recherche en Santé [CERS], Ministère de la Santé, Ministère de l’Enseignement Supérieur, de la Recherche Scientifique et de l’Innovation, Burkina Faso, No. 2020-4-076, dated 8 April 2020; Ethics Committee for Research on Human Beings of the Faculty of Medicine of São José do Rio Preto, FAMERP, Brazil, No. 4.032.814, dated 18 May 2020 for the prospective collection and No. 02078812.8.0000.5415, dated 27 May 2019 for the retrospective collection). Purchased samples were collected and approved for use for research purposes by the respective commercial providers (Boca Biolistics, Discovery Life Sciences and Atreide Biosamples). All participants, or a parent or legal guardian in case of children, gave informed consent before the start of the study.

Precision experiments were conducted using characterised negative and positive samples (bioMérieux collection). Cross-reactivity experiments were performed using samples collected from patients with other potentially interfering infections (bioMérieux collection) or from contrived samples generated from characterised negative samples. Negative samples were provided by the French blood bank (Etablissement Français du Sang [EFS], La Plaine Saint-Denis, France). Each volunteer donor signed a written informed consent for the use of blood for research purposes. EFS obtained from the French ministry of research the authorization to collect and transfer samples to partners (Ministère de l’Enseignement Supérieur, de la Recherche et de l’Innovation, reference AC-2017-2958).

### 2.2. Study Design and Definitions

This multicenter study aimed to evaluate the diagnostic performance of VIDAS^®^ dengue assays (bioMérieux SA, Marcy-l’Étoile, France) detecting DENV NS1 antigen (VIDAS^®^ DENGUE NS1 Ag) and anti-DENV IgM and IgG antibodies (VIDAS^®^ Anti-DENGUE IgM and VIDAS^®^ Anti-DENGUE IgG, respectively), in terms of positive percent agreement (PPA) and negative percent agreement (NPA) with competitor ELISA used as comparative method (Table 2).

**Table 2.** Competitor ELISA used for the concordance analysis with the VIDAS^®^ Dengue assays

| Competitor ELISA | Name of assay | Provider |
| --- | --- | --- |
| NS1 <sup>1</sup> | Dengue NS1 Antigen DxSelect™ (EL1510; IfU Rev. C) | Focus Diagnostics, DiaSorin Molecular LLC, Cypress, CA, USA |
|  | DENV Detect™ NS1 ELISA (DNS1-1; IfU No. 900109-02) | InBios International, Inc., Seattle, Washington, USA |
| IgM <sup>2</sup> | Panbio Dengue IgM Capture ELISA (01PE20) | Abbott Laboratories, Abbott Park, IL, USA |
|  | DENV Detect IgM Capture ELISA (DDMS-1) | InBios International, Inc., Seattle, Washington, USA |
| IgG | Panbio Dengue IgG Indirect ELISA (01PE30) | Abbott Laboratories, Abbott Park, IL, USA |

<sup>1</sup> Both NS1 ELISA were considered as equivalent based on respective instructions for use (IfU) and thus used interchangeably. <sup>2</sup> The results of both IgM ELISA were used to interpret the competitor IgM ELISA results (considered positive when both were positive, negative when both were negative, and undetermined when at least one was discordant or equivocal).

Quantitative RT-PCR and serotyping assays detecting all four DENV serotypes’ RNA were performed using the CE-marked VIASURE Dengue Virus Real Time PCR Detection Kit (CerTest Biotec, Zaragoza, Spain; site 1), the CDC DENV TaqMan^®^ RT-qPCR assay (Center for Disease Control and Prevention, USA; site 2; [25,26]), and the CE-marked RealStar Dengue RT-PCR (altona diagnostics GmbH, Hamburg, Germany; samples of site 3, outsourced to BIOMEX GmbH, Heidelberg, Germany).

Patient samples were split as per algorithm, based on competitor ELISA and RT-PCR results. The dengue NS1, IgM and IgG competitor ELISA test results, together with DENV RNA measurement by RT-PCR, were used to define the stage of DENV infection (Table 3). Samples positive for DENV RNA and/or NS1 antigen were defined as Acute DENV infection, regardless of the IgM and/or IgG ELISA results. Samples negative for both DENV RNA and NS1 antigen and positive for both IgM and IgG ELISA were defined as Post-Acute DENV infection. Samples negative for DENV RNA, NS1 antigen and IgM ELISA, but positive for IgG ELISA were defined as Recovery stage. Samples negative for all assays (RT-PCR and competitor ELISA) were defined as naïve for DENV (i.e., from patients with other febrile illnesses). Samples that did not meet these criteria or yielded undetermined test results were labelled as Unclassified (Table 3).

**Table 3.** Definition of DENV infection stages according to the competitor ELISA and RT-PCR test results.

| Stage of DENV infection | RT-PCR and/or NS1 competitor ELISA result <sup>1</sup> | IgM competitor ELISA results <sup>2</sup> | IgG competitor ELISA result <sup>3</sup> |
| --- | --- | --- | --- |
| Naïve <sup>8</sup> | Negative <sup>4</sup> | Negative | Negative |
| Acute | Positive <sup>5</sup> | Positive or negative | Positive or negative |
| Post-Acute | Negative <sup>4</sup> | Positive | Positive |
| Recovery | Negative <sup>4</sup> | Negative | Positive |
| Unclassified | Negative <sup>4</sup> or Undetermined <sup>6</sup> | Positive or Undetermined <sup>7</sup> | Negative or Undetermined <sup>6</sup> |

<sup>1</sup> NS1 ELISA as described in Table 2. <sup>2</sup> IgM ELISA as described in Table 2; both assays must be concordant (negative or positive); discordant competitor IgM ELISA results were excluded from the analysis (see Figure 1). <sup>3</sup> IgG ELISA as described in Table 2. <sup>4</sup> Negative for both RT-PCR and NS1 ELISA. <sup>5</sup> Positive for RT-PCR and/or NS1 ELISA. <sup>6</sup> Undetermined defined as equivocal ELISA (according to the manufacturer's definition). <sup>7</sup> Undetermined defined as either equivocal or discordant IgM ELISA (Panbio® Dengue IgM Capture ELISA and InBios DENV Detect IgM Capture ELISA). <sup>8</sup> Defines patients enrolled with a suspected DENV infection but were negative in all dengue-specific assays (RT-PCR and competitor ELISA) and are thus likely to suffer from another febrile illness.

Unclassified samples were excluded from analyses considering the stage of infection but were included in analyses based on the global population (Figure 1). Samples tested with all assays (VIDAS^®^ dengue assays, competitor ELISA and RT-PCR) were included in the final analysis (Figure 1).

### 2.3. VIDAS^®^ Assays

The VIDAS^®^ DENGUE NS1 Ag (DEAG; 423077), VIDAS^®^ Anti-DENGUE IgM (DENM; 423078) and VIDAS^®^ Anti-DENGUE IgG (DENG; 423079) assays (bioMérieux SA, Marcy-l’Étoile, France) are automated qualitative two-step immunoassays developed for VIDAS^®^ instruments [24]. The VIDAS^®^ DENGUE NS1 Ag assay detects the dengue NS1 antigen of the four DENV serotypes. The VIDAS^®^ Anti-DENGUE IgM and IgG assays detect IgM and IgG antibodies, respectively, recognising antigens of the four DENV serotypes, owing to the use of a recombinant tetravalent EDIIIT2 protein composed of the antigenic DENV-specific envelope domain III of the four DENV serotypes [24,27]. The three VIDAS^®^ dengue assays were performed and interpreted according to the instructions for use, as previously described [24]. A test was interpreted as negative when the index value (i) was < 1.0 and positive when i ≥ 1.0. VIDAS^®^ assays do not have equivocal test results.

### 2.4. Competitor Assays

Competitor ELISA (Table 2) were conducted and interpreted according to the manufacturers’ recommendations. NS1 ELISA (FOCUS, InBios), IgM ELISA (Panbio) and IgG ELISA (Panbio) were interpreted as negative for result values (index or immune status ratio [ISR]) < 0.9, positive for values > 1.1 and equivocal for 0.9–1.1 result values. IgM ELISA (InBios) was interpreted as negative for ISR ≤ 1.65, positive for ISR ≥ 2.84 and equivocal for 1.65–2.84 ISR values. While one NS1 ELISA (either FOCUS EL1510, Rev. C or InBios DNS1-1, version 900228-01, both considered equivalent according to their respective package insert) and one IgG ELISA (Panbio) were used as reference for the comparative analysis, both IgM ELISA (Panbio and InBios) were performed and considered for the competitor test evaluation. Thus, the competitor IgM ELISA was interpreted as negative when both IgM ELISA (Panbio and InBios) were negative, positive when both IgM ELISA were positive and undetermined when the Panbio and InBios IgM ELISA were discordant or equivocal (Table 3). Samples with undetermined and equivocal competitor ELISA test results for a particular marker were excluded from the concordance analysis for that marker (Figure 1).

### 2.5. Precision Experiments

Assay precision was evaluated according to the Clinical and Laboratory Standards Institute (CLSI) EP05-A3 guideline [28], using characterised negative and positive (native or contrived) human sera that were aliquoted and had undergone one freeze-thaw cycle. Precision experiments were conducted at the Clinical Affairs Laboratory, bioMérieux (Marcy l’Etoile, France). Within-run precision (repeatability) and within-laboratory precision (between-lot reproducibility) of the VIDAS^®^ Dengue NS1 Ag, Anti-IgM and Anti-IgG assays was determined using three samples each: one (high) negative and two (low and moderate) positive. Samples were run in duplicate on one VIDAS^®^ instrument, twice a day over 10 days (with an instrument calibration every second day), using two assay lots, thus generating 80 measurement values per sample (240 measurement values per VIDAS^®^ Dengue assay). A visual data integrity check was performed to identify possible outliers. Visually discordant results were confirmed to be statistical outliers using the Generalized Extreme Studentized Deviate (ESD) test with a 1% α risk. In case of confirmed outlier, the test was repeated, and the valid result was used for precision calculation only if a definite root cause was identified. Variance was expressed as standard deviation (SD) and coefficient of variation (CV).

### 2.6. Cross-reactivity Experiments

Cross-reactivity experiments were performed in adherence to the CLSI EP07-Ed3 guideline [29] at the Clinical Affairs Laboratory, bioMérieux (Marcy l’Etoile, France) using either contrived samples or native samples from patients with other potentially interfering infections tested positive either for the respective pathogen or for the respective pathogen-specific antigen, RNA, IgM, IgG or total antibodies.

Contrived samples were generated for evaluating cross-reactivity with the VIDAS^®^ Dengue NS1 Ag assay, by spiking defined amounts (10, 25, 50 ng/ml final concentration) of commercially available recombinant or inactivated native antigens (The Native Antigen Company, Oxford, UK; Meridian Bioscience, Cincinnati, Ohio, USA) into samples tested negative with the competitor NS1 ELISA (InBios DENV Detect™ NS1 ELISA or FOCUS DENGUE NS1 Antigen DxSelect™). PCR- or antigen-positive HBV, HCV and SARS-CoV-2 samples were used to test the cross-reactivity with the VIDAS^®^ Dengue NS1 Ag assay.

Samples used for evaluating the cross-reactivity with VIDAS^®^ Anti-Dengue IgM were positive for pathogen-specific IgM, except for IAV/IBV and HIV, which were positive for pathogen-specific total antibodies, HCV samples, which were positive for either anti-HCV IgG or anti-HCV total antibodies, ZIKV samples, which were positive for either anti-ZIKV IgM or anti-ZIKV IgG antibodies, and *Plasmodium falciparum* samples which were positive for the pathogen itself. Samples used for evaluating the cross-reactivity with VIDAS^®^ Anti-Dengue IgG were positive for pathogen-specific IgG, except for IAV/IBV, HBV and HIV, which were positive for pathogen-specific total antibodies, HCV samples, which were positive for either anti-HCV IgG or anti-HCV total antibodies, ZIKV samples, which were positive for either anti-ZIKV IgM or anti-ZIKV IgG antibodies, Yellow Fever samples, which were defined by the detection of neutralising antibodies, and *Plasmodium falciparum* samples, which were positive for the pathogen itself.

The dengue-negative status of all samples was determined using the respective competitor ELISA: NS1 ELISA for cross-reactivity with VIDAS^®^ Dengue NS1 Ag, Panbio Dengue IgM ELISA for cross-reactivity with VIDAS^®^ Anti-Dengue IgM, and Panbio Dengue IgG ELISA for cross-reactivity with VIDAS^®^ Anti-Dengue IgG.

Cross-reactivities were tested in singlicate using three lot each of the VIDAS^®^ Dengue NS1 Ag, Anti-Dengue IgM and Anti-Dengue IgG assays, on three instruments of the VIDAS^®^ family. In case of a positive test result, the assay was repeated in duplicate on one VIDAS^®^ instrument. A total of 68, 259 and 167 samples with other potentially interfering infections were tested on the VIDAS^®^ Dengue NS1 Ag, VIDAS^®^ Anti-Dengue IgM, and VIDAS^®^ Anti-Dengue IgG assays, respectively.

### 2.7. Statistical Analyses

Assay precision was assessed in adherence to the CLSI EP05-A3 guideline [28] by a component-of-variance analysis for nested design (Restricted Maximum Likelihood) using the SAS Enterprise Guide 7.12 software.

Concordance analyses were conducted between the VIDAS^®^ assays and competitor ELISA used as comparative method. Accordingly, the terms “positive percent agreement” (PPA) and “negative percent agreement” (NPA) were used instead of “sensitivity” and “specificity”, respectively. Concordance analyses (PPA, NPA and overall percent agreement) were performed in adherence to the CLSI EP12-A2 guideline [30]. The 95% confidence intervals (95% CI) were computed, either as Wilson Score Confidence Interval if the percentage agreement was in the range] 5%, 95%[or as Exact Binomial Confidence Interval otherwise, using the SAS Enterprise Guide 7.12 software.

Positive agreement between the dengue NS1 assays (whether VIDAS^®^ or competitor ELISA) and RT-PCR, corresponding to assay sensitivity relative to RT-PCR set as gold standard, was also evaluated, and the respective 95% CI were computed as above. The sensitivity of both DENV NS1 assays were compared by calculating the Cohen’s kappa coefficient (κ); a κ = 0.81 – 0.99 was interpreted as near-perfect agreement between the two NS1 assays, indicating that the difference in sensitivity to RT-PCR is not statistically significant.

VIDAS^®^ dengue index values were displayed as Tukey box plots, in the global population and according to the DENV infection stage defined by our algorithm (Table 3), using GraphPad Prism 5.04 (GraphPad Software, San Diego, CA, USA).

## 3. Results

### 3.1. Patients’ Characteristics

Out of 1636 patients with suspected DENV infection recruited at multiple dengue-endemic regions worldwide, 1296 eligible and tested with all assays (VIDAS^®^ assays, competitor ELISA and RT-PCR) were included in the analysis (Figure 1). 1205/1296 (93.0%) samples were assigned a DENV infection stage (Table 4) based on the test results of RT-PCR and the competitor ELISA (Table 3) and were included in the analysis per DENV infection stage (Figure 1).

**Table 4.** Patients’ characteristics.

|  |  |
| --- | --- |
| Study population, *N* (%) | 1296 (100.0%) |
| Study population according to the testing site, *N* (%) |  |
| Site 1 (Burkina Faso; sample collection and testing) | 480 (37.0%) |
| Site 2 (Brazil; sample collection and testing) | 392 (30.3%) |
| Site 3 (France; externally acquired sample testing) | 424 (32.7%) |
| Age in years, median (range) | 32.0 (5–88) |
| Sex, *N* (%) |  |
| Female | 741 (57.2%) |
| Male | 555 (42.8%) |
| RT-PCR-positive samples, *N* (%) | 154 (11.9%) |
| Serotype distribution among RT-PCR-positive samples, *N* (%) |  |
| DENV-1 | 74/154 (48.0%) |
| DENV-2 | 72/154 (46.8%) |
| DENV-3 | 6/154 (3.9%) |
| DENV-4 | 2/154 (1.3%) |
| Sample distribution according to time from symptom onset, *N* (%) |  |
| 0 – 3 days | 477 (36.8%) |
| 4 – 5 days | 228 (17.6%) |
| 6 – 8 days | 456 (35.2%) |
| 9 – 15 days | 78 (6.0%) |
| 16 days – 1 month | 34 (2.6%) |
| > 1 month | 16 (1.2%) |
| Unknown | 7 (0.6%) |
| DENV infection stage <sup>1</sup>, *N* (%) |  |
| Acute | 281 (21.7%) |
| Post-Acute | 123 (9.5%) |
| Recovery | 626 (48.3%) |
| Naïve | 175 (13.5%) |
| Unclassified | 91 (7.0%) |

<sup>1</sup> According to the RT-PCR- and competitor ELISA-based algorithm (Table 3).

Between 392 and 480 samples were tested at each of the three testing sites (Table 4). Included patients were mainly adults (1258/1296 [97.1%] ≥ 18 years-old), with a median age of 32 years, and 57.2% were female (Table 4). Among the 154 patients with a positive dengue-specific RT-PCR test result, most were of the DENV-1 (74/154 [48.0%]) and DENV-2 (72/154 [46.8%]) serotype (Table 4). The distribution of patients according to the time post symptom onset at inclusion is shown for the global population (Table 4) and for each DENV infection stage (Table S1). As expected, most (258/281 [91.8%]) acute samples (NS1 ELISA- and/or RT-PCR-positive) had a documented time from symptom onset of 0–8 days and most (101/123 [82.1%]) post-acute samples (NS1 ELISA- and RT-PCR-negative, IgM and IgG ELISA-positive) of 6– 8 days (Table S1). The observation that most (546/626 [87.2%]) recovery samples (NS1 ELISA- and RT-PCR-negative, IgM ELISA-negative, IgG ELISA-positive) and most (160/175 [91.4%]) naïve samples (negative for RT-PCR and all three dengue ELISA) according to our algorithm were documented with an early time from symptom onset (0–8 days; Table S1) indicate that these patients suffer from a febrile illness other than DENV.

### 3.2. VIDAS^®^ Dengue Test Result Description

Index distribution of the VIDAS^®^ Dengue NS1 Ag, Anti-dengue IgM and Anti-dengue IgG assays is shown in Figure S1, and the respective index medians and interquartile ranges are presented in Table S2. In line with the algorithm-based classification (Table 3), higher VIDAS^®^ Dengue NS1 Ag index values were observed in acute samples (Figure S1). VIDAS^®^ Dengue NS1 Ag index values were also higher when considering the samples collected within the first five days post symptom onset (D0-5) compared to the whole acute category. Similarly, VIDAS^®^ Anti-dengue IgM index values were highest in acute and post-acute samples, and VIDAS^®^ Anti-dengue IgG index values were highest in acute, post-acute and recovery samples (Figure S1).

In agreement with these index value results, evaluation of the qualitative VIDAS^®^ Dengue test results showed a high proportion (80.4%) of positive VIDAS^®^ Dengue NS1 Ag tests in acute samples within the first five days after symptom onset (D0-5), and a moderate to high proportion of positive VIDAS^®^ Anti-dengue IgM and Anti-dengue IgG tests in post-acute samples (74.8% and 80.5%, respectively) (Table S3). Interestingly, the proportion of positive VIDAS^®^ NS1 and/or IgM combined tests (NS1/IgM) raised to 86.5% in D0-5 acute samples and that of positive VIDAS^®^ IgM and/or IgG combined tests (IgM/IgG) raised to 91.9% in post-acute samples (Table S3).

### 3.3. Analytical Performance of the VIDAS^®^ Dengue Assays

#### 3.3.1. Assay Precision

Assay precision of the three VIDAS^®^ Dengue NS1 Ag, Anti-dengue IgM and Anti-dengue IgG assays was evaluated on negative and positive samples. The coefficient of variation (CV) did not exceed 7.5% for repeatability (within-run precision) and 10.7% for within-laboratory (between-lot) precision, across the three VIDAS^®^ assays (Table 5).

**Table 5.**
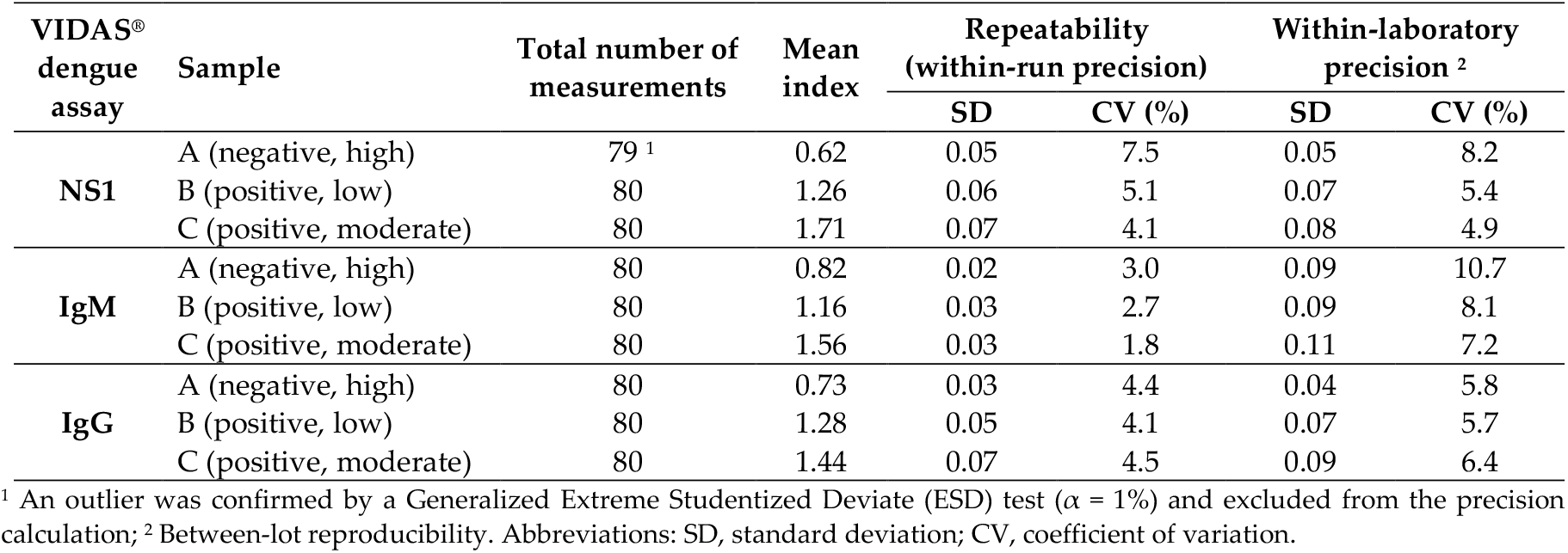
Precision of the VIDAS^®^ Dengue NS1, IgM and IgG assays.

| VIDAS®<br>dengue<br>assay | Sample | Total number of<br>measurements | Mean<br>index | Repeatability<br>(within-run precision) |  | Within-laboratory<br>precision <sup>2</sup> |  |
| --- | --- | --- | --- | --- | --- | --- | --- |
|  |  |  |  | SD | CV (%) | SD | CV (%) |
| NS1 | A (negative, high) | 79 <sup>1</sup> | 0.62 | 0.05 | 7.5 | 0.05 | 8.2 |
|  | B (positive, low) | 80 | 1.26 | 0.06 | 5.1 | 0.07 | 5.4 |
|  | C (positive, moderate) | 80 | 1.71 | 0.07 | 4.1 | 0.08 | 4.9 |
| IgM | A (negative, high) | 80 | 0.82 | 0.02 | 3.0 | 0.09 | 10.7 |
|  | B (positive, low) | 80 | 1.16 | 0.03 | 2.7 | 0.09 | 8.1 |
|  | C (positive, moderate) | 80 | 1.56 | 0.03 | 1.8 | 0.11 | 7.2 |
| IgG | A (negative, high) | 80 | 0.73 | 0.03 | 4.4 | 0.04 | 5.8 |
|  | B (positive, low) | 80 | 1.28 | 0.05 | 4.1 | 0.07 | 5.7 |
|  | C (positive, moderate) | 80 | 1.44 | 0.07 | 4.5 | 0.09 | 6.4 |

<sup>1</sup> An outlier was confirmed by a Generalized Extreme Studentized Deviate (ESD) test ( $\alpha = 1\%$ ) and excluded from the precision calculation; <sup>2</sup> Between-lot reproducibility. Abbreviations: SD, standard deviation; CV, coefficient of variation.

#### 3.3.2. Assay Cross-reactivity

Analytical specificity of the three VIDAS^®^ Dengue assays was evaluated using samples from patients with other proven infections and confirmed negative with the respective dengue ELISA reference tests. Cross-reactivity was measured as the proportion of positive VIDAS^®^ Dengue NS1, IgM and IgG assays (Table 6).

**Table 6.** Cross-reactivity of human contrived samples or native samples from patients with other infections potentially interfering with the VIDAS^®^ dengue NS1, IgM and IgG assays.

| Potentially interfering infections | Proportion of cross-reactions with VIDAS®<br>Dengue assays |  |  |
| --- | --- | --- | --- |
|  | NS1 | IgM | IgG |
| Herpes simplex virus (HSV1/2) | 0/6 <sup>1</sup> | 2/13 | 3/14 |
| Varicella zoster virus (VZV) | 0/3 <sup>1</sup> | 4/15 | 0/10 |
| Cytomegalovirus (CMV) | 0/3 <sup>1</sup> | 4/17 | 0/10 |
| Epstein-Barr virus (EBV) | 0/3 <sup>1</sup> | 3/14 | 2/15 |
| Influenza virus (IAV/IBV) | nd | 2/11 | 2/10 |
| *Borrelia burgdorferi* | nd | 2/14 | nd |
| *Plasmodium falciparum* | 0/3 <sup>1</sup> | 3/10 | 0/10 |
| Leptospira | nd | 3/12 | nd |
| Chikungunya virus (CHIKV) | 0/3 <sup>1</sup> | 0/13 | 0/7 |
| West Nile virus (WNV) | 0/3 <sup>1</sup> | 1/18 | 2/9 |
| Yellow fever virus (YFV) | 0/3 <sup>1</sup> | 2/21 | 0/14 |
| Zika virus (ZIKV) | 0/3 <sup>1</sup> | 1/14 | 0/8 |
| Hepatitis A virus (HAV) | nd | 1/15 | nd |
| Hepatitis B virus (HBV) | 0/10 | 2/14 | 0/7 |
| Hepatitis C Virus (HCV) | 1/9 | 3/14 | 1/14 |
| Parvovirus B19 | nd | 4/14 | 0/10 |
| Human immunodeficiency virus (HIV) | nd | 1/14 | 2/9 |
| HIV P24 antigen | 0/9 | nd | nd |
| Severe acute respiratory syndrome coronavirus 2<br>(SARS-CoV-2) | 0/10 | 2/16 | 1/20 |
| Total (%) | 1/68 (1.5%) | 40/259 (15.4%) | 13/167 (7.8%) |

<sup>1</sup> Contrived samples generated using volunteer donor samples tested negative with the competitor NS1 ELISA and spiked-in with 10, 25 or 50 ng/ml recombinant or inactivated native antigens. Abbreviation: nd, not determined.

Overall, cross-reactivity with the VIDAS^®^ Dengue NS1 Ag assay was very low (1/68 [1.5%]), with one reactivity with a native sample positive for hepatitis C virus (HCV) and none with samples of patients infected with severe acute respiratory syndrome coronavirus 2 (SARS-CoV-2) or with other flaviviruses such as West Nile virus, yellow fever virus and Zika virus (Table 6). Cross-reactivity with the VIDAS^®^ Anti-dengue IgG assay was higher (13/167 [7.8%]) and involved mainly samples of patients positive for IgG or total antibodies directed against other viruses (HSV, EBV, influenza, HCV, HIV), including the flavivirus West Nile virus (2/9) and the coronavirus SARS-CoV-2 (1/20) (Table 6). Crossreactivity with the VIDAS^®^ Anti-dengue IgM assay was the highest (40/259 [15.4%]). It involved samples of patients positive for antibodies against other viruses (VZV, CMV, EBV, HCV, parvovirus B19), bacteria (leptospira) and parasites (*Plasmodium falciparum*), and to some extent against the flaviviruses West Nile virus (1/18), yellow fever virus (2/21) and Zika virus (1/14), as well as the coronavirus SARS-CoV-2 (2/16) (Table 6).

### 3.4. Clinical Performance of the VIDAS^®^ Dengue Assays

#### 3.4.1. Clinical Sensitivity

Quantitative RT-PCR is often used as gold standard for the detection of acute DENV infection [20,24,31]. To evaluate the sensitivity of the VIDAS^®^ Dengue NS1 Ag assay, its positive agreement with DENV-specific RT-PCR set as gold standard was evaluated in acute samples and compared to that of the competitor NS1 ELISA (Table 7). No statistically significant differences were observed between the clinical sensitivity of VIDAS^®^ DENGUE NS1 Ag assay and that of the NS1 ELISA, on all acute samples (Cohen’s kappa coefficient κ = 0.885, *p* < 0.0001) and on acute samples collected within the first five days post onset of symptoms (D0-5) (Cohen’s kappa coefficient κ = 0.867, *p* < 0.0001). Sensitivity of the VIDAS^®^ Dengue NS1 Ag assay was higher in the D0-5 acute samples (81/106 [76.4%]) compared to all acute samples (98/153 [64.1%]) (Table 7).

**Table 7.** Positive agreement of the VIDAS^®^ and competitor ELISA NS1 antigen assays with RT-PCR.

| Reference test | Population | VIDAS® NS1 |  | NS1 ELISA <sup>2</sup> |  |
| --- | --- | --- | --- | --- | --- |
|  |  | *n/N* <sup>1</sup> | % [95% CI] | *n/N* <sup>1</sup> | % [95% CI] |
| RT-PCR | All acute samples | 98/153 | 64.1% [56.2-71.2] | 102/153 | 66.7% [58.9-73.6] |
|  | D0-5 acute samples | 81/106 | 76.4% [67.5-83.5] | 82/106 | 77.4% [68.5-84.3] |

<sup>1</sup> *n/N* is the ratio of the number of samples positive for the respective immunoassays (VIDAS® NS1, NS1 ELISA) to the number of RT-PCR-positive samples. To calculate sensitivity on the same cohort, one sample equivocal with the NS1 ELISA was excluded from the analysis. <sup>2</sup> InBios DENV Detect™ NS1 ELISA or FOCUS DENGUE NS1 Antigen DxSelect™. VIDAS® DENGUE NS1 Ag assay and NS1 ELISA showed a near-perfect agreement in their sensitivity to RT-PCR, with Cohen's kappa coefficients $\kappa = 0.885$ (all acute samples) and $\kappa = 0.867$ (D0-5 acute samples) ( $p < 0.0001$ in both cases). Abbreviations: CI, confidence interval; D0-5, 0-5 days post symptom onset.

#### 3.4.2. Clinical Specificity

To estimate the true-negative rate (specificity) of the three VIDAS^®^ Dengue assays, their negative agreement with the respective competitor ELISA was calculated in the population of naïve patients. Naïve patients are defined as subjects presenting clinical symptoms consistent with a DENV infection (i.e., the intended population of the VIDAS^®^ Dengue assays) but tested negative by RT-PCR and for the three markers (NS1, IgM and IgG) by the competitor ELISA (Table 3). The negative agreement of the VIDAS^®^ Dengue assays with the competitor ELISA was high in the population naïve for DENV, with 152/175 (86.9%) for VIDAS^®^ Anti-dengue IgM, 156/175 (89.1%) for VIDAS^®^ Anti-dengue IgG, and 175/175 (100.0%) for VIDAS^®^ Dengue NS1 Ag (Table 8).

**Table 8.**
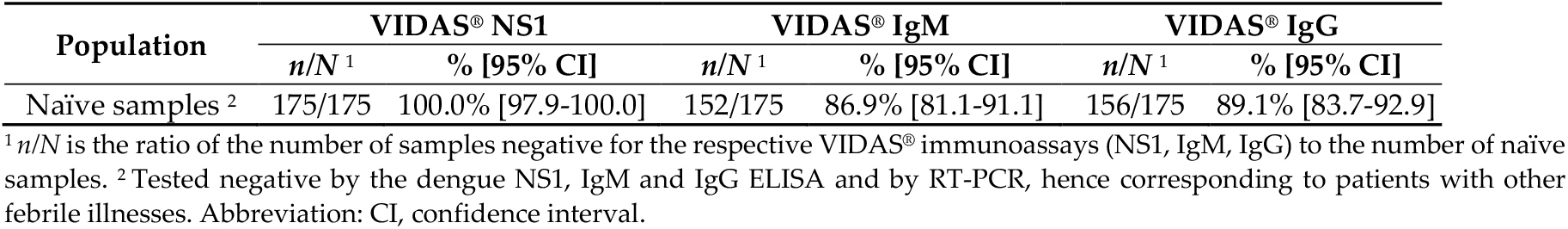
Negative agreement of the VIDAS^®^ Dengue assays with the competitor ELISA tested on study samples defined as naïve.

| Population | VIDAS® NS1 |  | VIDAS® IgM |  | VIDAS® IgG |  |
| --- | --- | --- | --- | --- | --- | --- |
|  | *n/N* <sup>1</sup> | % [95% CI] | *n/N* <sup>1</sup> | % [95% CI] | *n/N* <sup>1</sup> | % [95% CI] |
| Naïve samples <sup>2</sup> | 175/175 | 100.0% [97.9-100.0] | 152/175 | 86.9% [81.1-91.1] | 156/175 | 89.1% [83.7-92.9] |

<sup>1</sup> *n/N* is the ratio of the number of samples negative for the respective VIDAS® immunoassays (NS1, IgM, IgG) to the number of naïve samples. <sup>2</sup> Tested negative by the dengue NS1, IgM and IgG ELISA and by RT-PCR, hence corresponding to patients with other febrile illnesses. Abbreviation: CI, confidence interval.

#### 3.4.3. Concordance of DENV Infection Classification

Overall agreement of the DENV infection classification by the VIDAS^®^ Dengue assays with that by the competitor ELISA (as defined using the algorithm presented in Table 3), is shown in Table 9. Acute classification by the VIDAS^®^ Dengue test results was in strong agreement with the classification by the competitor ELISA, with up to 156/163 (95.7%) concordant classification for the early (D0-5) acute stage (Table 9). Classification of naïve samples was also highly concordant between VIDAS^®^ and competitor ELISA (142/175 [81.1%]) (Table 9). Concordance of post-acute (78/123 [63.4%]) and recovery (430/626 [68.7%]) classifications by VIDAS^®^ and competitor ELISA was moderate (Table 9). Since these stages are defined by IgM and/or IgG test positivity, partial concordance suggests differences in IgM and/or IgG test results between VIDAS^®^ and competitor ELISA. This is supported by the observation that discordant post-acute and recovery classifications were mainly classified in the respective category (i.e., most discordant post-acute were classified as recovery and vice versa by VIDAS^®^) (Table S4). Concordance of unclassified samples was very low (2/91 [2.2%]; Table 9). This was expected given that equivocal and undetermined test interpretations by the competitor ELISA does not occur with the VIDAS^®^ assays (which are interpreted as either positive or negative).

**Table 9.** Concordance of the infection stage classification by the VIDAS^®^-based *vs*. the competitor ELISA-based algorithm

| Stage of DENV infection <sup>2</sup> | Agreement |  |
| --- | --- | --- |
|  | *n*/*N* <sup>1</sup> | % [95% CI] |
| Acute (all) | 249/281 | 88.6% [84.4-91.8] |
| Acute (D0-5) | 156/163 | 95.7% [91.4-98.3] |
| Post-Acute | 78/123 | 63.4% [54.6-71.4] |
| Recovery | 430/626 | 68.7% [65.0-72.2] |
| Naïve | 142/175 | 81.1% [74.7-86.2] |
| Unclassified | 2/91 | 2.2% [0.3-7.7] |

<sup>1</sup> *n*/*N* is the ratio of the number of samples whose classification using the defined rules (Table 3) by the VIDAS® assays is concordant with that by the competitor ELISA, to the number of samples classified by the competitor ELISA. <sup>2</sup> According to the RT-PCR- and competitor ELISA-based algorithm (Table 3). Abbreviations: CI, confidence interval; D0-5, 0-5 days post symptom onset.

#### 3.4.4. Assay Concordance in the Total Study Population and per DENV Infection Stage

We next evaluated the positive (PPA) and negative (NPA) percent agreement of the three VIDAS^®^ dengue assays with the respective competitor ELISA (Table 10).

**Table 10.**
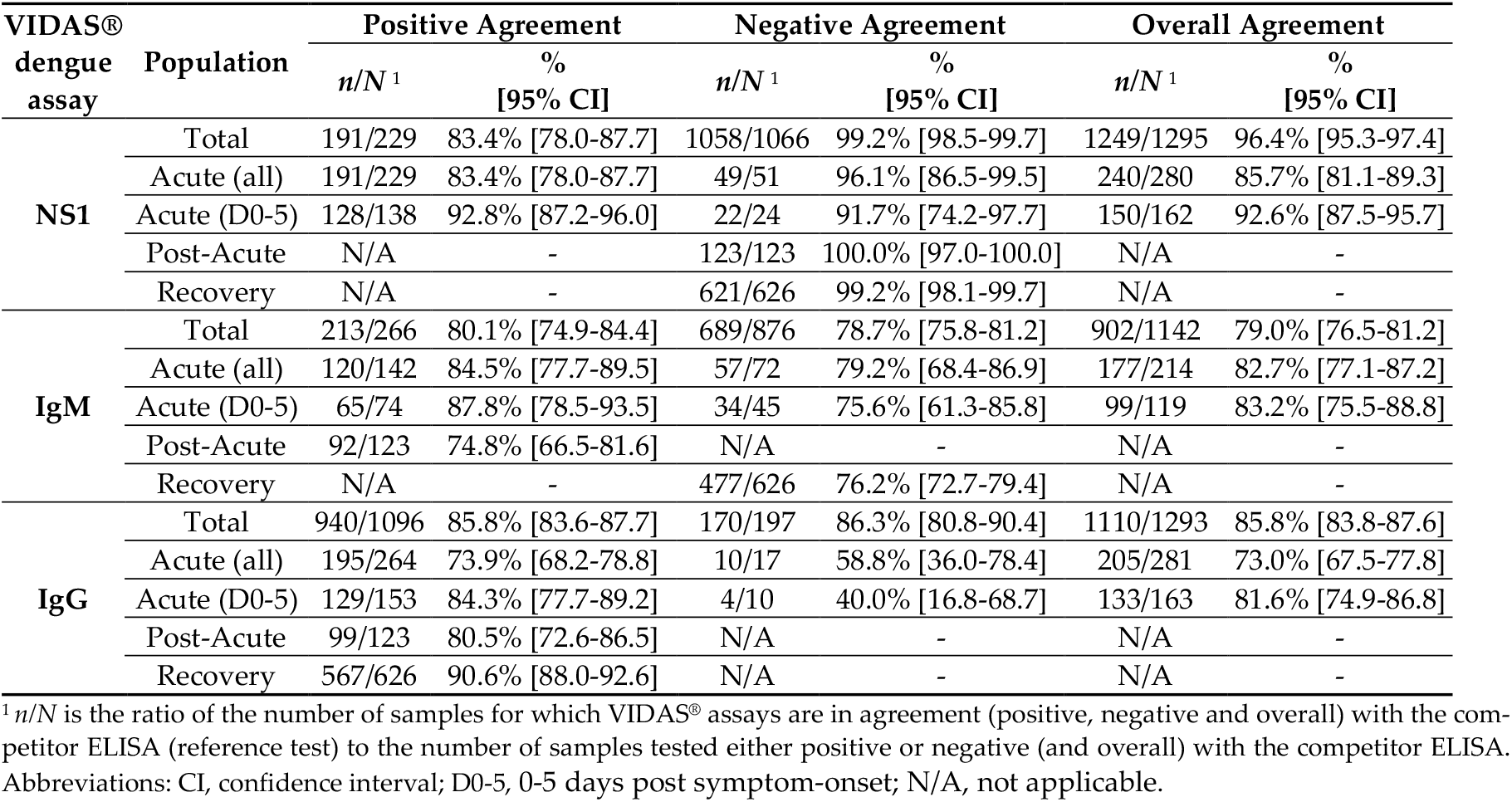
Concordance of the VIDAS^®^ dengue assays with the respective competitor ELISA, in the global population and per stage of DENV infection.

The PPA (95% CI) of the VIDAS^®^ Dengue NS1 Ag assay was 83.4% (78.0-87.7) in all acute samples, reaching 92.8% (87.2-96.0) in the D0-5 acute population (Table 10). The respective NPA (95% CI) was 99.2% (98.5-99.7) in the total population, ranging from 91.7% (74.2-97.7) in D0-5 acute to 100.0% (97.0-100.0) in post-acute samples (Table 10).

The PPA and NPA of the VIDAS^®^ Anti-dengue IgM assay were overall moderate, with a PPA (95% CI) of 80.1% (74.9-84.4) and a NPA (95% CI) of 78.7% (75.8-81.2) in the total population (Table 10).

The PPA (95% CI) of the VIDAS^®^ Anti-dengue IgG assay was 85.8% (83.6-87.7) in the total population, ranging from 73.9% (68.2-78.8) in acute to 90.6% (88.0-92.6) in recovery samples. Its NPA (95% CI) was 86.3% (80.8-90.4) in the total population, with a minimum of 40.0% (16.8-68.7) in D0-5 acute samples (Table 10).

Concordance analyses based on the cohort common to the three VIDAS^®^ dengue assays (Figure S2) yielded comparable results (Table S5). Moreover, concordance analyses according to time post symptom onset (instead of infection stage, as defined by our algorithm), revealed comparable results (Table S6). Notably, the PPA of the NS1 assay was highest (>90%) in the 0 – 5 days post symptom onset, and decreased afterward, as expected for this acute-phase marker and in line with current guidelines [5,7,8].

#### 3.4.5. Positive Agreement of Combined NS1/IgM and IgM/IgG Tests

Combining NS1, IgM and IgG test results is known to improve both sensitivity and specificity of DENV infection diagnosis, especially when patients present at different time points after symptom onset [5,7–10,13,14,22,24]. Thus, we evaluated the positive agreement of combined VIDAS^®^ NS1/IgM and IgM/IgG test results in the total population and at relevant infection stages, i.e., acute stage for NS1/IgM and post-acute stage for IgM/IgG (Table 11).

**Table 11.** Positive agreement of combined NS1/IgM and IgM/IgG VIDAS^®^ dengue assays with the respective competitor ELISA, in the total, acute (NS1/IgM) and post-acute (IgM/IgG) stage of DENV infection.

| VIDAS® dengue assay | Population | Positive Agreement |  |
| --- | --- | --- | --- |
|  |  | *n/N*<sup>1</sup> | % [95% CI] |
| NS1/IgM<sup>2</sup> | Total | 316/362 | 87.3% [83.5-90.3] |
|  | Total (D0-5) | 145/152 | 95.4% [90.7-98.1] |
|  | Acute (all) | 223/238 | 93.7% [89.9-96.1] |
|  | Acute (D0-5) | 134/139 | 96.4% [91.8-98.8] |
| IgM/IgG<sup>3</sup> | Total | 1011/1100 | 91.9% [90.1-93.4] |
|  | Post-Acute | 113/123 | 91.9% [85.7-95.5] |

<sup>1</sup> *n/N* is the ratio of the number of samples positive for the indicated combined VIDAS® Dengue assays (NS1- and/or IgM-positive; IgM- and/or IgG-positive) to the number of samples positive for the combined competitor ELISA. <sup>2</sup> PPA of VIDAS® NS1 and/or IgM *vs.* competitor NS1 and/or IgM ELISA; <sup>3</sup> PPA of VIDAS® IgM and/or IgG *vs.* competitor IgM and/or IgG ELISA. Abbreviations: CI, confidence interval; D0-5, 0-5 days post symptom-onset.

PPA (95% CI) of combined NS1/IgM in the total population was 87.3% (83.5-90.3) (Table 11) *vs*. 83.4% (78.0-87.7) for NS1 and 80.1% (74.9-84.4) for IgM (Table 10). PPA (95% CI) of combined NS1/IgM in acute samples was 93.7% (89.9-96.1), reaching 96.4% (91.8-98.8) in D0-5 acute samples (Table 11) (*vs*. 92.8% [87.2-96.0] for NS1 and 87.8% [78.5-93.5] for IgM; Table 10).

PPA (95% CI) of combined IgM/IgG in the total population was 91.9% (90.1-93.4) (Table 11) *vs*. 80.1% (74.9-84.4) for IgM and 85.8% (83.6-87.7) for IgG (Table 10). The PPA (95% CI) of combined IgM/IgG in post-acute samples was 91.9% (85.7-95.5) (Table 11) *vs*. 74.8% (66.5-81.6) for IgM and 80.5% (72.6-86.5) for IgG (Table 10).

## 4. Discussion

This multicenter study evaluated the diagnostic performance of three VIDAS^®^ dengue immunoassays detecting DENV NS1 antigen and Anti-DENV IgM and IgG antibodies, in comparison with commercial competitor ELISA. This study is a follow-up of the recent performance evaluation study conducted in 91 Lao patients with acute dengue infection, using the respective VIDAS^®^ dengue prototype assays [24]. A major strength of this study is the inclusion of a large number of samples (*n* = 1296) covering most dengue-endemic regions around the world, including Asia (India, Vietnam, The Philippines), Latin America (Brazil, Peru, Honduras, Dominican Republic) and Africa (Burkina Faso) [2,3].

The three VIDAS^®^ dengue assays demonstrated high within-run and within-laboratory precision (CV < 11.0%). The VIDAS^®^ DENGUE NS1 Ag assay showed very low cross-reactivity (1.5%) with samples positive for other infection-specific antigens. The VIDAS^®^ Anti-DENGUE IgM and IgG assays presented higher cross-reactivity (15.4% and 7.8%, respectively) with samples positive for IgM or IgG directed against other pathogen-specific antigens, as previously described for immunoassays [5,6,16,18,22,32–36]. These included cross-reactions with antibodies directed against other flaviviruses (4/53 [7.5%] for IgM and 2/31 [6.5%] for IgG). These cross-reactivity rates were however in range with or lower than those reported for other flavivirus IgM and IgG assays [34], including some of the competitor ELISA used in this study. Notably, cross-reactivity of the VIDAS^®^ Anti-DENGUE IgM and IgG assays with West Nile Virus (WNV)-positive samples was lower than that reported by the competitor IgM and IgG ELISA manufacturers (1/18 [5.6%] for VIDAS^®^ Anti-DENGUE IgM *vs*. 2/10 [20.0%] and 12/24 [50.0%] for IgM ELISA; 2/9 [22.2%] for VIDAS^®^ Anti-DENGUE IgG *vs*. 2/5 [40.0%] for IgG ELISA). Cross-reactivity with samples from SARS-CoV-2-infected patients (2/16 [12.5%] for IgM and 1/20 [5.0%] for IgG) was also observed. Such cross-reactions were expected, in line with the recent demonstration of shared antigenic similarity between DENV and SARS-CoV-2, resulting in cross-reactivity of the antibody response to both viruses [37–44]. Cross-reactivity of antibodies against DENV and SARS-CoV-2 was shown to be especially high using rapid diagnostic tests (RDT), as opposed to ELISA [37,39–44], in line with the relatively low cross-reactivity rates with SARS-CoV-2 samples observed in this study using the VIDAS^®^ Anti-DENV IgM and IgG assays.

Such antigenic similarity and cross-reactivity of immune responses of DENV and SARS-CoV-2 raise new diagnostic challenges in DENV-endemic regions in this time of COVID-19 pandemics [38,42,45–47]. Indeed, the impact of antibody cross-reactivity and/or of co-infection with DENV and SARS-CoV-2 on misdiagnosis but also on the immune response of patients and consequently on the outcome of immunoassays, remains unexplored. Given the partial overlap of sample collection with the COVID-19 pandemics in our study, and without documented information on possible co-infections with SARS-CoV-2, the results presented here should be interpreted with caution, as a possible impact of the COVID-19 pandemics cannot be excluded. Future studies should specifically explore this question.

Clinical performance of the VIDAS^®^ Dengue assays was assessed by measuring the sensitivity of the VIDAS^®^ DEN-GUE NS1 Ag assay (positive agreement with RT-PCR set as gold standard), and by evaluating assay concordance (positive and negative agreement with competitor ELISA). Sensitivity of the VIDAS^®^ DENGUE NS1 Ag assay in the D0-5 acute population (76.4%) was similar to that observed in the adult Lao population (79.5%) at a comparable time from symptom onset (median time of 4.5 days) [24]. Specificity of the three VIDAS^®^ Dengue assays (negative agreement with the respective competitor ELISA) in the intended population (i.e., patients with suspected DENV infection but classified as naïve for DENV) was also comparable to that measured in an adult healthy population using the three VIDAS^®^ Dengue prototypes (86.9–100.0% *vs*. 96.1–100.0%, respectively) [24].

Overall, concordance analyses revealed a good agreement of the VIDAS^®^ Dengue assays with competitor ELISA. As to DENV infection classification, the strong agreement (95.7%) of D0-5 acute stage classification by VIDAS^®^ Dengue assays using our staging algorithm is in line with the high PPA of the VIDAS^®^ DENGUE NS1 Ag assay in the D0-5 acute population (92.8%) and the comparable sensitivity of the VIDAS^®^ DENGUE NS1 Ag and competitor NS1 ELISA (76.4% *vs*. 77.4%) toward RT-PCR. The NPA of the VIDAS^®^ DENGUE NS1 Ag assay was also very high, at all DENV infection stages (91.7%–100.0%). Of note, in the adult acute Lao population, the PPA of the VIDAS^®^ DENGUE NS1 Ag prototype was slightly higher (98.0%) and the NPA slightly lower (87.5%) than in the current multicenter study [24]. This might be in part due to the difference in sample number between both studies.

On the other hand, the moderate agreement of post-acute (63.4%) and recovery (68.7%) stage classification by VIDAS^®^ Dengue assays is in line with the partial concordance with competitor ELISA of the VIDAS^®^ Anti-DENGUE IgM assay (PPA of 74.8% in post-acute samples, NPA of 76.2% in recovery samples) and to some extent of the VIDAS^®^ Anti-DENGUE IgG assay (PPA of 80.5% in post-acute samples). These results indicate a higher discordance of IgM and/or IgG test results between VIDAS^®^ and competitor ELISA.

Assay concordance was improved in combined NS1/IgM test results at early (D0-5) acute stages of DENV infection (96.4% PPA), and in combined IgM/IgG test results post-acute (91.9% PPA), demonstrating the benefit of combining marker immunodetection using the VIDAS^®^ Dengue assays to improve the sensitivity of diagnosis of DENV infection, as previously reported for other assays [5,9,10,13,24].

This study presents several limitations. First, despite the large number of collected samples (392 to 480 per site), heterogeneity between testing sites were observed, notably in the distribution of infection stage, time post symptom onset, and serotype distribution. This precluded subgroup analyses per site. Second, 146/154 (94.8%) samples were of DENV-1 and DENV-2 serotypes. While DENV-1 and DENV-2 serotypes are predominant in most dengue-endemic regions worldwide [4], it is possible that our performance evaluation study is not representative of DENV-3 and DENV-4 serotypes. Given that all four DENV serotypes are known to co-circulate in a spatial and/or temporal manner [48], and that assay performance might be serotype-dependent [6], additional studies should be conducted to confirm assay performance in patients infected with DENV-3 and DENV-4 serotypes. Third, the choice of considering two competitor IgM ELISA as comparator to the VIDAS^®^ Anti-DENGUE IgM assay led to the exclusion of 152 samples from the IgM analysis (because discordant between the two competitor IgM ELISA), which might have introduced a bias in the analysis. Since the concordance analysis conducted on the common samples showed comparable performance results, such bias is however unlikely. Fourth, as mentioned above, we cannot exclude an effect of the COVID-19 pandemics (via cross-reactivity and/or co-infection interferences) on the VIDAS^®^ Dengue assay performance evaluation, although the impact might be limited in the context of a concordance analysis (assuming a similar impact on both compared assays). Finally, the use of distinct and non-standardized RT-PCR and serotyping methods at the three testing sites might have introduced a bias in the acute infection stage definition according to our algorithm, although this is unlikely given the high sensitivity and specificity (90–100%) of commercial RT-PCR assays [1,5,8,9,13].

## 5. Conclusions

Altogether, this multicenter study conducted on a large number of samples representative of several dengue-endemic regions demonstrated a strong performance of the VIDAS^®^ DENGUE NS1 Ag assay, either alone or in combination with the VIDAS^®^ Anti-DENGUE IgM assay, notably at the early stage of DENV infection (first five days post symptom onset), and a strong performance of the VIDAS^®^ Anti-DENGUE IgG assay at later stages of infection, either alone or in combination with the VIDAS^®^ Anti-DENGUE IgM assay. These results therefore further support the use of the three VIDAS^®^ Dengue immunoassays as a complete solution for the rapid, automated and reliable diagnosis of DENV infection in dengue-endemic regions.

## Data Availability

All data produced in the present work are contained in the manuscript.

## Supplementary Materials

The following are available online at www.mdpi.com/xxx/s1, Figure S1: Index values of the VIDAS^®^ dengue assays, in the total population and per DENV infection stage, Figure S2: Extended study flow diagram describing the concordance analysis on common samples shown in Table S5, Table S1: Distribution of samples according to the time from symptom onset, per DENV infection stage, Table S2: Median and interquartile range (IQR) of VIDAS^®^ dengue assays, in the total population and per DENV infection stage (see Figure S1), Table S3: Percentage of positive test results of combined NS1/IgM and IgM/IgG VIDAS^®^ dengue assays *vs*. single assays, in the acute and post-acute stages of DENV infection, respectively, Table S4: Description of the discordant infection stage classifications between the VIDAS^®^-based and the competitor ELISA-based algorithm, Table S5: Concordance of the VIDAS^®^ dengue assays with the respective competitor ELISA, in the global population and per stage of DENV infection (common samples; see Figure S2), Table S6: Concordance of the VIDAS^®^ dengue assays with the respective competitor ELISA, according to time intervals post symptom onset.

## Author Contributions

Conceptualization, M.T., S.D. and M.L.N.; methodology, L.B. and L.D.; software, L.D.; validation, M.T., S.D. and M.L.N.; formal analysis, L.B.; investigation, A.F.V., A.K., T.M.I.L.S., B.H.G.A.M., A.C., A.C.S. and F.S.; resources, A.K., C.F.E., A.C., A.C.S. and F.S.; data curation, L.D.; writing—review and editing, M.T., L.B., S.D., A.F.V. and M.L.N.; supervision, S.D. and M.L.N.; project administration, M.T. All authors have read and agreed to the published version of the manuscript.

## Institutional Review Board Statement

The study was conducted according to the guidelines of the Declaration of Helsinki, and approved by the respective institutional review boards (Comité d’Éthique pour la Recherche en Santé [CERS], Ministère de la Santé, Ministère de l’Enseignement Supérieur, de la Recherche Scientifique et de l’Innovation, Burkina Faso, No. 2020-4-076, dated 8 April 2020; Ethics Committee for Research on Human Beings of the Faculty of Medicine of São José do Rio Preto, FAMERP, Brazil, No. 4.032.814, dated 18 May 2020 for the prospective collection and No. 02078812.8.0000.5415, dated 27 May 2019 for the retrospective collection).

## Informed Consent Statement

Written informed consent was obtained from all subjects involved in the study.

## Data Availability Statement

The data presented in this study are available within the article and supplementary material.

## Acknowledgments

The authors are grateful to Agnès Foussadier (bioMérieux) for her scientific contribution, Françoise Gay-Andrieu (bioMérieux) for her medical advisory support, Nadège Goutagny (bioMérieux) for discussions on the manuscript, Nafissatou Ilboudo, Fatoumata Maïga, Zoumana Bado, Adéline Sandwidi, Félix Lompo and Yacouba Barro (IRSS) for their contribution in patient enrolment and baseline data collection, and to Jacques simpore (CERBA), Théophane Yonli (CERBA) and Arthur Djibougou (IRSS/Centre MURAZ) for their support in the conduct of the clinical trial. A.F.V. was a postdoctoral fellow of the São Paulo Research Foundation (FAPESP #2018/17647-0) during the time of this research. The authors thank Anne Rascle of AR Medical Writing (Regensburg, Germany) for providing medical writing support, which was funded by bioMérieux (Marcy L’Etoile, France), in accordance with Good Publication Practice (GPP3) guidelines (http://www.ismpp.org/gpp3; accessed 11 February 2022).

## Supplementary Materials

**Figure S1.**
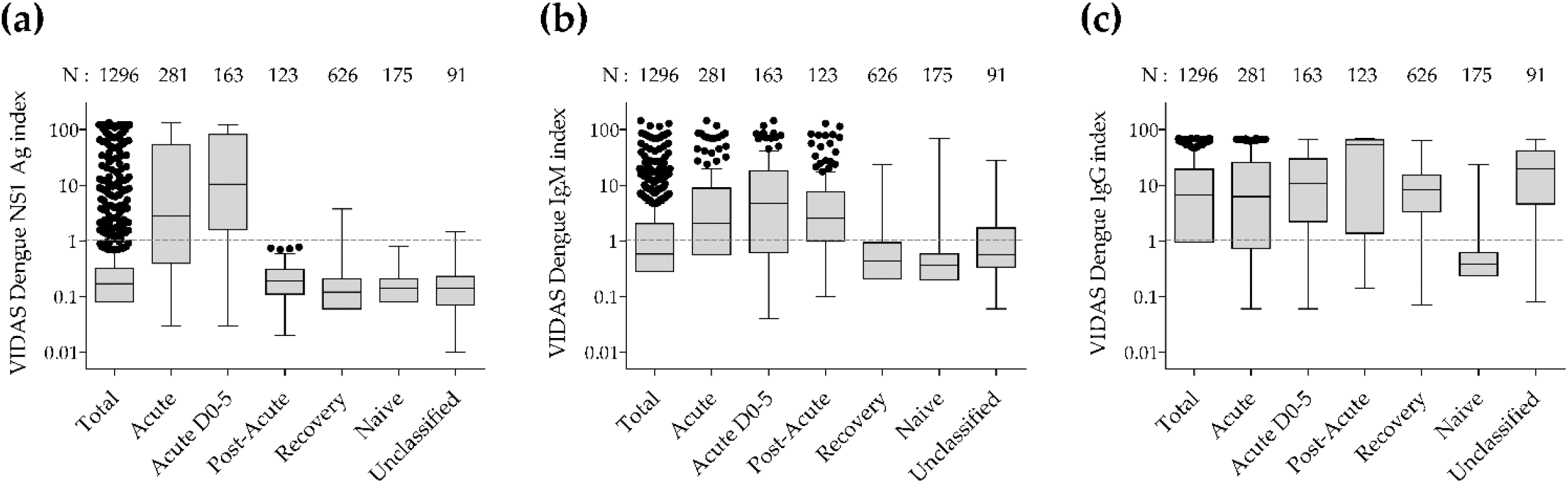
Index values of the VIDAS^®^ dengue assays, in the total population and per DENV infection stage. VIDAS^®^ Dengue NS1 Antigen (**a**), IgM (**b**) and IgG (**c**) assays were performed as described in the Methods section. Index values are depicted as Tukey boxplots in the whole study population (Total) and according to the DENV infection stages (Acute, Acute within 0 to 5 days post symptom onset [D0-5], Post-Acute, Recovery, Naïve and Unclassified), as defined in Table 3. The grey dashed line indicates the positivity cutoff (i = 1.0). The number of samples (N) per group is shown above each box. Respective median (interquartile range) index values are shown in Table S2.

**Figure S2.**
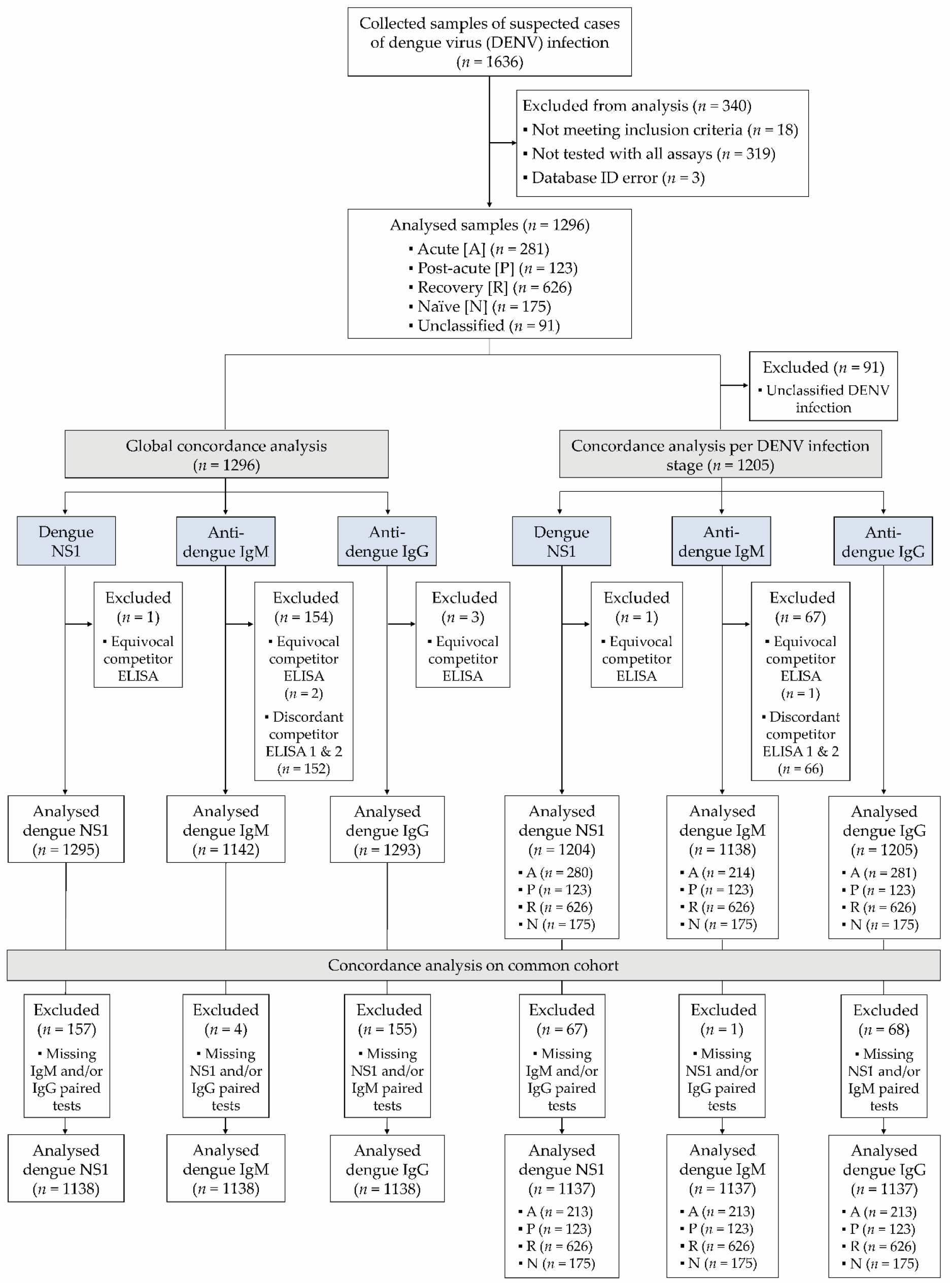
Extended study flow diagram describing the concordance analysis on common samples shown in Table S5.

**Table S1.** Distribution of samples according to the time from symptom onset, per DENV infection stage.

| Days from symptom onset | Stage of DENV infection <sup>1</sup>, *N* (%) |  |  |  |  | Total |
| --- | --- | --- | --- | --- | --- | --- |
|  | Acute | Post-Acute | Recovery | Naïve | Unclassified |  |
| **0 – 3 days** | 102 (36.3%) | 6 (4.9%) | 258 (41.2%) | 88 (50.3%) | 23 (25.3%) | 477 |
| **4 – 5 days** | 61 (21.7%) | 7 (5.7%) | 119 (19.0%) | 24 (13.7%) | 17 (18.7%) | 228 |
| **6 – 8 days** | 95 (33.8%) | 101 (82.1%) | 169 (27.0%) | 48 (27.4%) | 43 (47.2%) | 456 |
| **9 – 15 days** | 11 (3.9%) | 8 (6.5%) | 43 (6.9%) | 12 (6.9%) | 4 (4.4%) | 78 |
| **16 days – 1 month** | 7 (2.5%) | 0 (0.0%) | 23 (3.7%) | 2 (1.1%) | 2 (2.2%) | 34 |
| **> 1 month** | 1 (0.4%) | 0 (0.0%) | 12 (1.9%) | 1 (0.6%) | 2 (2.2%) | 16 |
| **Unknown** | 4 (1.4%) | 1 (0.8%) | 2 (0.3%) | 0 (0.0%) | 0 (0.0%) | 7 |
| **Total** | 281 | 123 | 626 | 175 | 91 | 1296 |

<sup>1</sup> As defined in Table 3.

**Table S2.** Median and interquartile range (IOR) of VIDAS^®^ dengue assays, in the total population and per DENV infection stage (see Figure S1).

| VIDAS®<br>dengue<br>assay | variable | DENV infection stage |  |  |  |  |  |  |
| --- | --- | --- | --- | --- | --- | --- | --- | --- |
|  |  | Total | Acute | Acute D0-5 | Post-Acute | Recovery | Naïve | Unclassified |
| NS1 | *N* | 1296 | 281 | 163 | 123 | 626 | 175 | 91 |
|  | Median | 0.2 | 2.8 | 10.5 | 0.2 | 0.1 | 0.1 | 0.1 |
|  | IQR | 0.1-0.3 | 0.4-54.1 | 1.6-81.1 | 0.1-0.3 | 0.1-0.2 | 0.1-0.2 | 0.1-0.2 |
| IgM | *N* | 1296 | 281 | 163 | 123 | 626 | 175 | 91 |
|  | Median | 0.6 | 2.1 | 4.7 | 2.5 | 0.4 | 0.4 | 0.6 |
|  | IQR | 0.3-2.0 | 0.6-8.9 | 0.6-18.0 | 1.0-7.6 | 0.2-0.9 | 0.2-0.6 | 0.3-1.7 |
| IgG | *N* | 1296 | 281 | 163 | 123 | 626 | 175 | 91 |
|  | Median | 6.7 | 6.2 | 10.7 | 53.0 | 8.2 | 0.4 | 19.6 |
|  | IQR | 1.0-19.2 | 0.7-25.3 | 2.2-29.7 | 1.4-65.3 | 3.4-15.1 | 0.2-0.6 | 4.6-41.5 |

**Table S3.** Percentage of positive test results of combined NSI/IgM and IgM/IgG VIDAS^®^ dengue assays *vs*. single assays, in the acute and post-acute stages of DENV infection, respectively.

| Population | VIDAS® dengue assay | % positive test results |  |
| --- | --- | --- | --- |
|  |  | *n*/*N* <sup>1</sup> | % [95% CI] |
| Acute (all) | NS1 | 194/281 | 69.0% [63.4-74.2] |
|  | IgM | 175/281 | 62.3% [56.5-67.7] |
|  | NS1/IgM <sup>2</sup> | 233/281 | 82.9% [78.1-86.9] |
| Acute (D0-5) | NS1 | 131/163 | 80.4% [73.6-85.7] |
|  | IgM | 108/163 | 66.3% [58.7-73.1] |
|  | NS1/IgM <sup>2</sup> | 141/163 | 86.5% [80.4-90.9] |
| Post-Acute | IgM | 92/123 | 74.8% [66.5-81.6] |
|  | IgG | 99/123 | 80.5% [72.6-86.5] |
|  | IgM/IgG <sup>3</sup> | 113/123 | 91.9% [85.7-95.5] |

<sup>1</sup> *n*/*N* is the ratio of the number of samples positive with the respective VIDAS® Dengue assays to the total number of samples in the indicated DENV infection stage. <sup>2</sup> Percentage of tests positive for VIDAS® NS1 and/or IgM; <sup>3</sup> Percentage of tests positive for VIDAS® IgM and/or IgG. Abbreviations: CI, confidence interval; D0-5, 0-5 days post symptom-onset.

**Table S4.** Description of the discordant infection stage classifications between the VIDAS^®^-based and the competitor ELISA-based algorithm

| Stage of DENV infection <sup>3</sup> | Disagreement | VIDAS®-based classification (*n*) <sup>2</sup> |  |  |  |  |
| --- | --- | --- | --- | --- | --- | --- |
|  | *n/N* <sup>1</sup> (%) | Acute | Post-Acute | Recovery | Naïve | Unclassified |
| **Acute (all)** | 32/281 (11.4%) | - | 18 | 4 | 3 | 7 |
| **Acute (D0-5)** | 7/163 (4.3%) | - | 5 | 1 | 0 | 1 |
| **Post-Acute** | 45/123 (36.6%) | 0 | - | 21 | 10 | 14 |
| **Recovery** | 196/626 (31.3%) | 5 | 132 | - | 46 | 13 |
| **Naïve** | 33/175 (18.9%) | 0 | 9 | 10 | - | 14 |
| **Unclassified** | 89/91 (97.8%) | 1 | 31 | 50 | 7 | - |

<sup>1</sup> *n/N* is the ratio of the number of samples whose classification using the defined rules (Table 3) by the VIDAS® assays was discordant with that by the competitor ELISA, to the number of samples classified by the competitor ELISA-based algorithm; <sup>2</sup> *n* is the number of discordant samples per stage (according to VIDAS®-based classification). <sup>3</sup> According to the RT-PCR- and competitor ELISA-based algorithm (Table 3).

**Table S5.** Concordance of the VIDAS^®^ dengue assays with the respective competitor ELISA, in the global population and per stage of DENV infection (common samples; see Figure S2)

| VIDAS®<br>dengue<br>assay | Population | Positive Agreement |  | Negative Agreement |  | Overall Agreement |  |
| --- | --- | --- | --- | --- | --- | --- | --- |
|  |  | *n/N*<sup>1</sup> | %<br>[95% CI] | *n/N*<sup>1</sup> | %<br>[95% CI] | *n/N*<sup>1</sup> | %<br>[95% CI] |
| NS1 | Total | 135/170 | 79.4% [72.7-84.8] | 963/968 | 99.5% [98.8-99.8] | 1098/1138 | 96.5% [95.2-97.5] |
|  | Acute (all) | 135/170 | 79.4% [72.7-84.8] | 43/43 | 100.0% [91.8-100.0] | 178/213 | 83.6% [78.0-87.9] |
|  | Acute (D0-5) | 89/98 | 90.8% [83.5-95.1] | 20/20 | 100.0% [83.2-100.0] | 109/118 | 92.4% [86.1-95.9] |
|  | Post-Acute | N/A | - | 123/123 | 100.0% [97.0-100.0] | N/A | - |
|  | Recovery | N/A | - | 621/626 | 99.2% [98.1-99.7] | N/A | - |
| IgM | Total | 213/266 | 80.1% [74.9-84.4] | 686/872 | 78.7% [75.8-81.3] | 899/1138 | 79.0% [76.5-81.3] |
|  | Acute (all) | 120/142 | 84.5% [77.7-89.5] | 57/71 | 80.3% [69.6-87.9] | 177/213 | 83.1% [77.5-87.5] |
|  | Acute (D0-5) | 65/74 | 87.8% [78.5-93.5] | 34/44 | 77.3% [63.0-87.2] | 99/118 | 83.9% [76.2-89.4] |
|  | Post-Acute | 92/123 | 74.8% [66.5-81.6] | N/A | - | N/A | - |
|  | Recovery | N/A | - | 477/626 | 76.2% [72.7-79.4] | N/A | - |
| IgG | Total | 811/948 | 85.5% [83.2-87.6] | 164/190 | 86.3% [80.7-90.5] | 975/1138 | 85.7% [83.5-87.6] |
|  | Acute (all) | 145/199 | 72.9% [66.3-78.6] | 8/14 | 57.1% [32.6-78.6] | 153/213 | 71.8% [65.4-77.4] |
|  | Acute (D0-5) | 91/110 | 82.7% [74.6-88.7] | 2/8 | 25.0% [7.1-59.1] | 93/118 | 78.8% [70.6-85.2] |
|  | Post-Acute | 99/123 | 80.5% [72.6-86.5] | N/A | - | N/A | - |
|  | Recovery | 567/626 | 90.6% [88.0-92.6] | N/A | - | N/A | - |

<sup>1</sup> *n/N* is the ratio of the number of samples for which VIDAS® assays are in agreement (positive, negative and overall) with the competitor ELISA (reference test) to the number of samples tested either positive or negative (and overall) with the competitor ELISA. Abbreviations: CI, confidence interval; D0-5, 0-5 days post symptom-onset; N/A, not applicable.

**Table S6.** Concordance of the VIDAS^®^ dengue assays with the respective competitor ELISA, according to time intervals post symptom onset.

|  |  | Positive Percent Agreement |  | Negative Percent Agreement |  | Overall Percent Agreement |  |
| --- | --- | --- | --- | --- | --- | --- | --- |
| VIDAS®<br>dengue<br>assay | Time interval post<br>symptom onset | *n*/*N*<sup>1</sup> | % [95% CI] | *n*/*N*<sup>1</sup> | % [95% CI] | *n*/*N*<sup>1</sup> | % [95% CI] |
| NS1 | 0 – 3 days | 79 / 84 | 94.05% [ 86.81 ; 97.43 ] % | 388 / 392 | 98.98% [ 97.41 ; 99.72 ] % | 467 / 476 | 98.11% [ 96.44 ; 99.13 ] % |
|  | 4 – 5 days | 49 / 54 | 90.74% [ 80.09 ; 95.98 ] % | 172 / 174 | 98.85% [ 95.91 ; 99.86 ] % | 221 / 228 | 96.93% [ 93.78 ; 98.76 ] % |
|  | 6 – 8 days | 55 / 77 | 71.43% [ 60.51 ; 80.31 ] % | 377 / 379 | 99.47% [ 98.11 ; 99.94 ] % | 432 / 456 | 94.74% [ 92.29 ; 96.44 ] % |
|  | 9 – 15 days | 4 / 7 | 57.14% [ 25.05 ; 84.18 ] % | 71 / 71 | 100.00% [ 94.94 ; 100.00 ] % | 75 / 78 | 96.15% [ 89.17 ; 99.20 ] % |
|  | 16 days – 1 month | 0 / 3 | 0.00% [ 0.00 ; 70.76 ] % | 31 / 31 | 100.00% [ 88.78 ; 100.00 ] % | 31 / 34 | 91.18% [ 77.04 ; 96.95 ] % |
|  | > 1 month | 0 / 0 | . [ 0.00 ; 0.00 ] % | 16 / 16 | 100.00% [ 79.41 ; 100.00 ] % | 16 / 16 | 100.00% [ 79.41 ; 100.00 ] % |
|  | Unknown | 4 / 4 | 100.00% [ 39.76 ; 100.00 ] % | 3 / 3 | 100.00% [ 29.24 ; 100.00 ] % | 7 / 7 | 100.00% [ 59.04 ; 100.00 ] % |
| IgM | 0 – 3 days | 39 / 46 | 84.78% [ 71.78 ; 92.43 ] % | 300 / 380 | 78.95% [ 74.57 ; 82.75 ] % | 339 / 426 | 79.58% [ 75.49 ; 83.13 ] % |
|  | 4 – 5 days | 37 / 41 | 90.24% [ 77.45 ; 96.14 ] % | 110 / 157 | 70.06% [ 62.49 ; 76.68 ] % | 147 / 198 | 74.24% [ 67.73 ; 79.83 ] % |
|  | 6 – 8 days | 126 / 164 | 76.83% [ 69.80 ; 82.63 ] % | 193 / 233 | 82.83% [ 77.47 ; 87.13 ] % | 319 / 397 | 80.35% [ 76.16 ; 83.96 ] % |
|  | 9 – 15 days | 9 / 11 | 81.82% [ 52.30 ; 94.86 ] % | 49 / 60 | 81.67% [ 70.08 ; 89.44 ] % | 58 / 71 | 81.69% [ 71.15 ; 88.98 ] % |
|  | 16 days – 1 month | 0 / 0 | . [ 0.00 ; 0.00 ] % | 25 / 31 | 80.65% [ 63.72 ; 90.81 ] % | 25 / 31 | 80.65% [ 63.72 ; 90.81 ] % |
|  | > 1 month | 1 / 1 | 100.00% [ 2.50 ; 100.00 ] % | 10 / 13 | 76.92% [ 49.74 ; 91.82 ] % | 11 / 14 | 78.57% [ 52.41 ; 92.43 ] % |
|  | Unknown | 1 / 3 | 33.33% [ 6.15 ; 79.23 ] % | 2 / 2 | 100.00% [ 15.81 ; 100.00 ] % | 3 / 5 | 60.00% [ 23.07 ; 88.24 ] % |
| IgG | 0 – 3 days | 344 / 380 | 90.53% [ 87.16 ; 93.08 ] % | 81 / 95 | 85.26% [ 76.77 ; 91.01 ] % | 425 / 475 | 89.47% [ 86.39 ; 91.92 ] % |
|  | 4 – 5 days | 174 / 200 | 87.00% [ 81.63 ; 90.97 ] % | 22 / 27 | 81.48% [ 63.30 ; 91.82 ] % | 196 / 227 | 86.34% [ 81.27 ; 90.21 ] % |
|  | 6 – 8 days | 326 / 400 | 81.50% [ 77.40 ; 85.00 ] % | 52 / 56 | 92.86% [ 83.02 ; 97.19 ] % | 378 / 456 | 82.89% [ 79.17 ; 86.07 ] % |
|  | 9 – 15 days | 52 / 66 | 78.79% [ 67.49 ; 86.92 ] % | 9 / 12 | 75.00% [ 46.77 ; 91.11 ] % | 61 / 78 | 78.21% [ 67.84 ; 85.92 ] % |
|  | 16 days – 1 month | 27 / 30 | 90.00% [ 74.38 ; 96.54 ] % | 4 / 4 | 100.00% [ 39.76 ; 100.00 ] % | 31 / 34 | 91.18% [ 77.04 ; 96.95 ] % |
|  | > 1 month | 11 / 14 | 78.57% [ 52.41 ; 92.43 ] % | 2 / 2 | 100.00% [ 15.81 ; 100.00 ] % | 13 / 16 | 81.25% [ 56.99 ; 93.41 ] % |
|  | Unknown | 6 / 6 | 100.00% [ 54.07 ; 100.00 ] % | 0 / 1 | 0.00% [ 0.00 ; 97.50 ] % | 6 / 7 | 85.71% [ 48.69 ; 97.43 ] % |

<sup>1</sup> *n*/*N* is the ratio of the number of samples for which VIDAS® assays are in agreement (positive, negative and overall) with the competitor ELISA (reference test) to the number of samples tested either positive or negative (and overall) with the competitor ELISA. Abbreviation: CI, confidence interval

## References

1. Harapan, H.; Michie, A.; Sasmono, R.T.; Imrie, A. Dengue: A Minireview. Viruses 2020, 12, 829, doi:10.3390/v12080829.

2. Wilder-Smith, A.; Ooi, E.-E.; Horstick, O.; Wills, B. Dengue. Lancet 2019, 393, 350–363, doi:10.1016/S0140-6736(18)32560-1.

3. Bhatt, S.; Gething, P.W.; Brady, O.J.; Messina, J.P.; Farlow, A.W.; Moyes, C.L.; Drake, J.M.; Brownstein, J.S.; Hoen, A.G.; Sankoh, O.; et al. The Global Distribution and Burden of Dengue. Nature 2013, 496, 504–507, doi:10.1038/nature12060.

4. Cucunawangsih, null; Lugito, N.P.H. Trends of Dengue Disease Epidemiology. Virology (Auckl) 2017, 8, 1178122X17695836, doi:10.1177/1178122X17695836.

5. Dengue: Guidelines for Diagnosis, Treatment, Prevention, and Control; WHO, Ed.; New edition.; World Health Organization (WHO) and the Special Programme for Research and Training in Tropical Diseases (TDR): Geneva, Switzerland, 2009; ISBN 978-92-4-154787-1.

6. Raafat, N.; Blacksell, S.D.; Maude, R.J. A Review of Dengue Diagnostics and Implications for Surveillance and Control. Transactions of The Royal Society of Tropical Medicine and Hygiene 2019, 113, 653–660, doi:10.1093/trstmh/trz068.

7. Center for Disease Control and Prevention Testing for Dengue Virus | Dengue | CDC Available online: https://www.cdc.gov/dengue/healthcare-providers/testing/index.html (accessed on 27 October 2021).

8. Pan American Health Organization; World Health Organization Dengue: Guidelines for Patient Care in the Region of the Americas; 2016; ISBN 978-92-75-31890-4.

9. Peeling, R.W.; Artsob, H.; Pelegrino, J.L.; Buchy, P.; Cardosa, M.J.; Devi, S.; Enria, D.A.; Farrar, J.; Gubler, D.J.; Guzman, M.G.; et al. Evaluation of Diagnostic Tests: Dengue. Nat Rev Microbiol 2010, 8, S30–38, doi:10.1038/nrmicro2459.

10. Blacksell, S.D.; Jarman, R.G.; Gibbons, R.V.; Tanganuchitcharnchai, A.; Mammen, M.P.; Nisalak, A.; Kalayanarooj, S.; Bailey, M.S.; Premaratna, R.; de Silva, H.J.; et al. Comparison of Seven Commercial Antigen and Antibody Enzyme-Linked Immunosorbent Assays for Detection of Acute Dengue Infection. Clin Vaccine Immunol 2012, 19, 804–810, doi:10.1128/CVI.05717-11.

11. Blacksell, S.D. Commercial Dengue Rapid Diagnostic Tests for Point-of-Care Application: Recent Evaluations and Future Needs? J Biomed Biotechnol 2012, 2012, 151967, doi:10.1155/2012/151967.

12. Blacksell, S.D.; Jarman, R.G.; Bailey, M.S.; Tanganuchitcharnchai, A.; Jenjaroen, K.; Gibbons, R.V.; Paris, D.H.; Premaratna, R.; de Silva, H.J.; Lalloo, D.G.; et al. Evaluation of Six Commercial Point-of-Care Tests for Diagnosis of Acute Dengue Infections: The Need for Combining NS1 Antigen and IgM/IgG Antibody Detection to Achieve Acceptable Levels of Accuracy. Clin Vaccine Immunol 2011, 18, 2095–2101, doi:10.1128/CVI.05285-11.

13. Muller, D.A.; Depelsenaire, A.C.I.; Young, P.R. Clinical and Laboratory Diagnosis of Dengue Virus Infection. J Infect Dis 2017, 215, S89–S95, doi:10.1093/infdis/jiw649.

14. Fry, S.R.; Meyer, M.; Semple, M.G.; Simmons, C.P.; Sekaran, S.D.; Huang, J.X.; McElnea, C.; Huang, C.-Y.; Valks, A.; Young, P.R.; et al. The Diagnostic Sensitivity of Dengue Rapid Test Assays Is Significantly Enhanced by Using a Combined Antigen and Antibody Testing Approach. PLoS Negl Trop Dis 2011, 5, e1199, doi:10.1371/journal.pntd.0001199.

15. Pal, S.; Dauner, A.L.; Valks, A.; Forshey, B.M.; Long, K.C.; Thaisomboonsuk, B.; Sierra, G.; Picos, V.; Talmage, S.; Morrison, A.C.; et al. Multicountry Prospective Clinical Evaluation of Two Enzyme-Linked Immunosorbent Assays and Two Rapid Diagnostic Tests for Diagnosing Dengue Fever. J Clin Microbiol 2015, 53, 1092–1102, doi:10.1128/JCM.03042-14.

16. Blessmann, J.; Winkelmann, Y.; Keoviengkhone, L.; Sopraseuth, V.; Kann, S.; Hansen, J.; El Halas, H.; Emmerich, P.; Schmidt-Chanasit, J.; Schmitz, H.; et al. Assessment of Diagnostic and Analytic Performance of the SD Bioline Dengue Duo Test for Dengue Virus (DENV) Infections in an Endemic Area (Savannakhet Province, Lao People’s Democratic Republic). PLoS One 2020, 15, e0230337, doi:10.1371/journal.pone.0230337.

17. Simonnet, C.; Okandze, A.; Matheus, S.; Djossou, F.; Nacher, M.; Mahamat, A. Prospective Evaluation of the SD BIOLINE Dengue Duo Rapid Test during a Dengue Virus Epidemic. Eur J Clin Microbiol Infect Dis 2017, 36, 2441–2447, doi:10.1007/s10096-017-3083-8.

18. Hunsperger, E.A.; Yoksan, S.; Buchy, P.; Nguyen, V.C.; Sekaran, S.D.; Enria, D.A.; Pelegrino, J.L.; Vázquez, S.; Artsob, H.; Drebot, M.; et al. Evaluation of Commercially Available Anti-Dengue Virus Immunoglobulin M Tests. Emerg Infect Dis 2009, 15, 436–440, doi:10.3201/eid1503.080923.

19. Pal, S.; Dauner, A.L.; Mitra, I.; Forshey, B.M.; Garcia, P.; Morrison, A.C.; Halsey, E.S.; Kochel, T.J.; Wu, S.-J.L. Evaluation of Dengue NS1 Antigen Rapid Tests and ELISA Kits Using Clinical Samples. PLoS One 2014, 9, e113411, doi:10.1371/journal.pone.0113411.

20. Gaikwad, S.; Sawant, S.S.; Shastri, J.S. Comparison of Nonstructural Protein-1 Antigen Detection by Rapid and Enzyme-Linked Immunosorbent Assay Test and Its Correlation with Polymerase Chain Reaction for Early Diagnosis of Dengue. J Lab Physicians 2017, 9, 177–181, doi:10.4103/0974-2727.208265.

21. Lee, H.; Ryu, J.H.; Park, H.S.; Park, K.H.; Bae, H.; Yun, S.; Choi, A.R.; Cho, S.Y.; Park, C.; Lee, D.G.; et al. Comparison of Six Commercial Diagnostic Tests for the Detection of Dengue Virus Non-Structural-1 Antigen and IgM/IgG Antibodies. Ann Lab Med 2019, 39, 566–571, doi:10.3343/alm.2019.39.6.566.

22. Hunsperger, E.A.; Yoksan, S.; Buchy, P.; Nguyen, V.C.; Sekaran, S.D.; Enria, D.A.; Vazquez, S.; Cartozian, E.; Pelegrino, J.L.; Artsob, H.; et al. Evaluation of Commercially Available Diagnostic Tests for the Detection of Dengue Virus NS1 Antigen and Anti-Dengue Virus IgM Antibody. PLoS Negl Trop Dis 2014, 8, e3171, doi:10.1371/journal.pntd.0003171.

23. Zhang, H.; Li, W.; Wang, J.; Peng, H.; Che, X.; Chen, X.; Zhou, Y. NS1-Based Tests with Diagnostic Utility for Confirming Dengue Infection: A Meta-Analysis. Int J Infect Dis 2014, 26, 57–66, doi:10.1016/j.ijid.2014.02.002.

24. Somlor, S.; Brossault, L.; Grandadam, M. Evaluation of VIDAS® Diagnostic Assay Prototypes Detecting Dengue Virus NS1 Antigen and Anti-Dengue Virus IgM and IgG Antibodies. Diagnostics (Basel) 2021, 11, 1228, doi:10.3390/diagnostics11071228.

25. Colombo, T.E.; Versiani, A.F.; Dutra, K.R.; Rubiato, J.G.D.; Galvão, T.M.; Negri Reis, A.F.; Nogueira, M.L. Performance of CDC Trioplex QPCR during a Dengue Outbreak in Brazil. J Clin Virol 2019, 121, 104208, doi:10.1016/j.jcv.2019.104208.

26. Johnson, B.W.; Russell, B.J.; Lanciotti, R.S. Serotype-Specific Detection of Dengue Viruses in a Fourplex Real-Time Reverse Transcriptase PCR Assay. J Clin Microbiol 2005, 43, 4977–4983, doi:10.1128/JCM.43.10.4977-4983.2005.

27. Combe, M.; Lacoux, X.; Martinez, J.; Méjan, O.; Luciani, F.; Daniel, S. Expression, Refolding and Bio-Structural Analysis of a Tetravalent Recombinant Dengue Envelope Domain III Protein for Serological Diagnosis. Protein Expr Purif 2017, 133, 57–65, doi:10.1016/j.pep.2017.03.004.

28. Clinical & Laboratory Standards Institute EP05-A3: Evaluating Quantitative Measurement Precision, 3rd Edition Available online: https://clsi.org/standards/products/method-evaluation/documents/ep05/ (accessed on 27 October 2021).

29. Clinical & Laboratory Standards Institute EP07: Interference Testing in Clinical Chemistry - Third Edition Available online: https://clsi.org/standards/products/method-evaluation/documents/ep07/ (accessed on 26 January 2022).

30. Clinical & Laboratory Standards Institute EP12-A2: User Protocol for Evaluation of Qualitative Test Performance, 2nd Edition Available online: https://clsi.org/standards/products/method-evaluation/documents/ep12/ (accessed on 12 October 2022).

31. Ambrose, J.H.; Sekaran, S.D.; Azizan, A. Dengue Virus NS1 Protein as a Diagnostic Marker: Commercially Available ELISA and Comparison to QRT-PCR and Serological Diagnostic Assays Currently Used by the State of Florida. J Trop Med 2017, 2017, 8072491, doi:10.1155/2017/8072491.

32. Tate, J.; Ward, G. Interferences in Immunoassay. Clin Biochem Rev 2004, 25, 105–120.

33. Piantadosi, A.; Kanjilal, S. Diagnostic Approach for Arboviral Infections in the United States. J Clin Microbiol 2020, 58, e01926–19, doi:10.1128/JCM.01926-19.

34. Endale, A.; Medhin, G.; Darfiro, K.; Kebede, N.; Legesse, M. Magnitude of Antibody Cross-Reactivity in Medically Important Mosquito-Borne Flaviviruses: A Systematic Review. Infect Drug Resist 2021, 14, 4291–4299, doi:10.2147/IDR.S336351.

35. Souza, N.C.S.E.; Félix, A.C.; de Paula, A.V.; Levi, J.E.; Pannuti, C.S.; Romano, C.M. Evaluation of Serological Cross-Reactivity between Yellow Fever and Other Flaviviruses. Int J Infect Dis 2019, 81, 4–5, doi:10.1016/j.ijid.2019.01.023.

36. Koraka, P.; Zeller, H.; Niedrig, M.; Osterhaus, A.D.M.E.; Groen, J. Reactivity of Serum Samples from Patients with a Flavivirus Infection Measured by Immunofluorescence Assay and ELISA. Microbes Infect 2002, 4, 1209–1215, doi:10.1016/s1286-4579(02)01647-7.

37. Lustig, Y.; Keler, S.; Kolodny, R.; Ben-Tal, N.; Atias-Varon, D.; Shlush, E.; Gerlic, M.; Munitz, A.; Doolman, R.; Asraf, K.; et al. Potential Antigenic Cross-Reactivity between SARS-CoV-2 and Dengue Viruses. Clinical Infectious Diseases 2020, ciaa1207, doi:10.1093/cid/ciaa1207.

38. Nath, H.; Mallick, A.; Roy, S.; Sukla, S.; Biswas, S. Computational Modelling Supports That Dengue Virus Envelope Antibodies Can Bind to SARS-CoV-2 Receptor Binding Sites: Is Pre-Exposure to Dengue Virus Protective against COVID-19 Severity? Comput Struct Biotechnol J 2021, 19, 459–466, doi:10.1016/j.csbj.2020.12.037.

39. Spinicci, M.; Bartoloni, A.; Mantella, A.; Zammarchi, L.; Rossolini, G.M.; Antonelli, A. Low Risk of Serological Cross-Reactivity between Dengue and COVID-19. Mem Inst Oswaldo Cruz 2020, 115, e200225, doi:10.1590/0074-02760200225.

40. Nath, H.; Mallick, A.; Roy, S.; Sukla, S.; Basu, K.; De, A.; Biswas, S. Archived Dengue Serum Samples Produced False-Positive Results in SARS-CoV-2 Lateral Flow-Based Rapid Antibody Tests. J Med Microbiol 2021, 70, doi:10.1099/jmm.0.001369.

41. Vanroye, F.; Bossche, D.V. den; Brosius, I.; Tack, B.; Esbroeck, M.V.; Jacobs, J. COVID-19 Antibody Detecting Rapid Diagnostic Tests Show High Cross-Reactivity When Challenged with Pre-Pandemic Malaria, Schistosomiasis and Dengue Samples. Diagnostics (Basel) 2021, 11, 1163, doi:10.3390/diagnostics11071163.

42. Masyeni, S.; Santoso, M.S.; Widyaningsih, P.D.; Asmara, D.W.; Nainu, F.; Harapan, H.; Sasmono, R.T. Serological Cross-Reaction and Coinfection of Dengue and COVID-19 in Asia: Experience from Indonesia. Int J Infect Dis 2021, 102, 152–154, doi:10.1016/j.ijid.2020.10.043.

43. Santoso, M.S.; Masyeni, S.; Haryanto, S.; Yohan, B.; Hibberd, M.L.; Sasmono, R.T. Assessment of Dengue and COVID-19 Antibody Rapid Diagnostic Tests Cross-Reactivity in Indonesia. Virol J 2021, 18, 54, doi:10.1186/s12985-021-01522-2.

44. Khairunisa, S.Q.; Amarullah, I.H.; Churrotin, S.; Fitria, A.L.; Amin, M.; Lusida, M.I.; Soegijanto, S. Potential Misdiagnosis between COVID-19 and Dengue Infection Using Rapid Serological Test. Infect Dis Rep 2021, 13, 540–551, doi:10.3390/idr13020050.

45. Harapan, H.; Ryan, M.; Yohan, B.; Abidin, R.S.; Nainu, F.; Rakib, A.; Jahan, I.; Emran, T.B.; Ullah, I.; Panta, K.; et al. Covid-19 and Dengue: Double Punches for Dengue-Endemic Countries in Asia. Rev Med Virol 2021, 31, e2161, doi:10.1002/rmv.2161.

46. Schulte, H.L.; Brito-Sousa, J.D.; Lacerda, M.V.G.; Naves, L.A.; de Gois, E.T.; Fernandes, M.S.; Lima, V.P.; Rassi, C.H.R.E.; de Siracusa, C.C.; Sasaki, L.M.P.; et al. SARS-CoV-2/DENV Co-Infection: A Series of Cases from the Federal District, Midwestern Brazil. BMC Infect Dis 2021, 21, 727, doi:10.1186/s12879-021-06456-2.

47. Mejía-Parra, J.L.; Aguilar-Martinez, S.; Fernández-Mogollón, J.L.; Luna, C.; Bonilla-Aldana, D.K.; Rodriguez-Morales, A.J.; Díaz-Vélez, C. Characteristics of Patients Coinfected with Severe Acute Respiratory Syndrome Coronavirus 2 and Dengue Virus, Lambayeque, Peru, May-August 2020: A Retrospective Analysis. Travel Med Infect Dis 2021, 43, 102132, doi:10.1016/j.tmaid.2021.102132.

48. Messina, J.P.; Brady, O.J.; Scott, T.W.; Zou, C.; Pigott, D.M.; Duda, K.A.; Bhatt, S.; Katzelnick, L.; Howes, R.E.; Battle, K.E.; et al. Global Spread of Dengue Virus Types: Mapping the 70 Year History. Trends Microbiol 2014, 22, 138–146, doi:10.1016/j.tim.2013.12.011.

